# A loss of function variant in *SLC30A8/ZnT8* drives proteomic changes associated with lowered apoptosis in human stem cell-derived islets

**DOI:** 10.64898/2026.04.17.26351108

**Authors:** Marie Gasser, Inès Cherkaoui, Giada Ostinelli, Mathieu Ferron, Qian Du, Dieter Egli, Guy A. Rutter

## Abstract

**Aims and hypothesis:** Loss-of-function mutations in *SLC30A8*, encoding the zinc ion (Zn^2+^) transporter ZnT8 in pancreatic beta cells, lower type 2 diabetes risk dose-dependently, but the underlying mechanisms remain unclear. Here, we combine proteomic, transcriptomic and functional approaches in human stem cell-derived islet-like clusters bearing common alleles or the inactivating variant R138X. We hypothesized that this variant protects against the deleterious effect of Zn^2+^ depletion on cell survival and function.

**Methods:** Human embryonic stem cells INS(GFP/w) (MEL1), and CRISPR/Cas9-derived heterozygous or homozygous R138X lines were differentiated into stem cell-derived islet-like clusters. Intracellular Zn^2+^ levels were reduced using the chelator N,N,N′,N′-tetrakis(2-pyridylmethyl)-1,2-ethanediamine (TPEN). Apoptosis was assessed by TUNEL staining and protein expression by immunofluorescence. Glucose-stimulated calcium (Ca^2+^) dynamics were measured using the intracellular probe (Cal590) and insulin secretion by homogenous time-resolved fluorescence. Transcriptomic profiling was performed by bulk mRNA sequencing and proteomics by liquid chromatography-tandem mass spectrometry.

**Results:** Intracellular Zn^2+^ depletion increased apoptosis in wild-type islet-like clusters, whereas R138X clusters were protected. R138X heterozygous clusters showed a mild increase in GCG^+^ cells and R138X homozygous clusters exhibited increased NKX6.1^+^ cells, without affecting polyhormonal populations. These changes were reversed under Zn^2+^ depletion. Transcriptomic and proteomic analyses, assessing genotype effects while accounting for Zn^2+^ depletion, showed that R138X clusters (versus wild-type) exhibited upregulation of genes and proteins involved in vesicle trafficking, secretion, Ca²⁺ signaling and mitochondrial metabolism, consistent with enhanced glucose-stimulated insulin secretion in homozygous clusters. Conversely, genes and proteins associated with extracellular matrix remodeling, metal-ion handling, apoptosis and cellular stress were downregulated. R138X clusters displayed altered Ca^2+^ signaling, with decreased area under the curve and oscillation amplitude, but increased frequency. These differences were reversed by TPEN, while Zn^2+^ depletion impaired Ca^2+^ response in wild-type clusters. Despite lowered overall activity, R138X homozygous clusters showed enhanced overall cell-cell connectivity, reversed by TPEN treatment. The opposite effects were observed in R138X heterozygous clusters, showing improved connectivity and activity under Zn^2+^ depletion.

**Conclusion and interpretation:** Intracellular Zn^2+^ depletion compromises islet-like cluster identity and function, while the R138X variant confers protection against these effects. Under Zn^2+^-depleted conditions, ZnT8 deficiency promotes a more mature and metabolically active state of the R138X clusters, with enhanced Ca^2+^ signaling and insulin secretion, supported by a structural remodeling and the downregulation of apoptosis and cellular stress. These findings highlight the therapeutic potential of targeting ZnT8 in type 2 diabetes and support its relevance for further improving cell-based therapies.

**Research in Context:** *What is already know about this subject?:*

- Rare inactivating mutations in the insulin granule-associated zinc transporter gene, *SLC30A8/ZnT8*, drive lowered type 2 diabetes risk.
- Previous studies have indicated that apoptosis is lowered, and glucose-stimulated insulin secretion enhanced, after ZnT8 inactivation.
- The molecular mechanisms underlying these changes are unclear.

*What is the key question?:*

- How do inactivating mutations in *SL30A8/ZnT8* lead to lowered apoptosis and enhanced insulin secretion from stem cell-derived islet-like clusters, and is altered susceptibility to intracellular zinc depletion involved?

*What are the new findings?:*

- The rare inactivating R138X mutation in *SLC30A8* leads to gene dose-dependent changes in the transcriptome and proteome of islet-like clusters.
- Changes include upregulation of maturity and downregulation of immaturity genes.
- Depletion of intracellular Zn^2+^ exaggerates the protective effects of the inactivating mutation on apoptosis and insulin secretion

*How might this impact on clinical practice in the foreseeable future?:*

- Our findings suggest that careful monitoring of both dietary zinc intake and of circulating levels of zinc ions, whose effects are mitigated in *SLC30A8* mutation carriers, may be helpful in some populations to lower diabetes risk.

## Introduction

Zinc is an essential trace element involved in primary metabolism, immune function and inflammation [1, 2]. Most importantly, zinc is an important regulator of apoptosis, required for cell survival. Zinc deficiency, due to decreased intake or malnutrition is a major health concern [3] and was defined by the WHO as a significant disease contributing factor. In line with this, alterations in intracellular zinc levels have been associated with obesity, type 2 diabetes and insulin resistance [4, 5]. In pancreatic islets, decreased zinc levels have been associated with beta cell dysfunction and death in mice [6].

Within beta cells, zinc transport is mediated by the zinc ion (Zn^2+^) transporter ZnT8 encoded by the *Solute Carrier Transporter Family 30 Member 8* (*SLC30A8*) gene, the most abundantly expressed Zn^2+^ transporter in beta cells. The mature protein is located on the membrane of insulin-secretory granules [7]. By facilitating Zn^2+^ influx into the granules, ZnT8 ensures subsequent insulin crystallization as insulin/zinc hexamers, the final step of insulin biosynthesis [7, 8].

Genetic studies have identified *SLC30A8* as a key locus associated with type 2 diabetes susceptibility. Common variants in the *SLC30A8* gene, including the non-synonymous single-nucleotide polymorphism R325W, are associated with a 10 to 20% increased risk of type 2 diabetes [9, 10]. This variant has been linked to impaired beta cell function and altered insulin secretion in both human and rodent models [11–13].

Paradoxically, the rare protein truncating variant of *SLC30A8*, pArg138* (R138X), confers protection against type 2 diabetes. Identified in 2014, heterozygous carriers of R138X exhibited a ∼60% reduced risk in developing type 2 diabetes [14], associated with improved insulin secretion, enhanced glucose responsiveness and more efficient proinsulin processing. More recently, a large-scale genetic study identified individuals with complete loss of *SLC30A8* and demonstrated a gene dose-dependent protection against type 2 diabetes, with odds ratios of ∼0.62 and ∼0.34 for heterozygous and homozygous carriers, respectively [15]. These findings were associated with lowered blood glucose levels, and enhanced beta cell function and insulin secretion, hence supporting the protective role of lowered ZnT8 activity in humans.

Consistent with these genetic observations, experimental models have shown that loss of ZnT8 function can confer protection to cellular stress. In mouse and human islets, as well as beta cell lines and stem cell-derived islet-like clusters, *SLC30A8* deletion or knockdown protect against the pro-apoptotic effects of zinc depletion, hypoxia, inflammatory cytokines and glucolipotoxicity [8, 16–20]. These protective effects were often associated with altered intracellular zinc distribution, within beta cells or across the whole pancreas [16, 19] and improved beta cell survival, although effects on insulin secretion vary according to the experimental model [21].

Despite these advances, the molecular mechanisms underlying the protective effect of reduced ZnT8 remain incompletely understood [22]. Here, we investigate the impact on the survival and function of human stem cell-derived islet-like clusters of the inactivating R138X *SLC30A8* variant, and explore how this is affected by intracellular Zn^2+^ depletion. We demonstrate that, in addition to the protection against apoptosis, R138X islets show a drastic transcriptomic and proteomic remodeling, leading to improved glucose-regulated Ca^2+^ dynamics and insulin secretion, changes exaggerated by Zn^2+^-depletion.

## Methods

Detailed methods are provided in Supplementary Methods.

### Differentiation of human embryonic stem cells

Human embryonic stem cells were obtained as described [8]. Cell lines with mutations were established by introducing heterozygous and homozygous p.Arg138* mutations into human embryonic stem cell line MEL1 using CRISPR/Cas9 [8] and differentiated according to [23].

### Zinc depletion

At the end of the differentiation, islet-like clusters were incubated with the Zn^2+^ chelator N,N,N′,N′-tetrakis(2-pyridinylmethyl)-1,2-ethanediamine (TPEN, Cat No. P4413, Sigma-Aldrich, USA) at 1 μM or vehicle for 48h at 37°C and 5% CO_2_ to induce intracellular Zn^2+^ depletion.

### Immunostaining

Islet-like clusters were fixed with 4% paraformaldehyde and cryosections were prepared as described previously [23].

### Proteomics

Protein lysates were extracted in lysis buffer and proteomic analysis was performed.

### RNA-Sequencing

Total mRNA was isolated and 500 ng of total RNA were used as input for bulk mRNA Sequencing (Illumina NovaSeq).

### TUNEL assay

Slides were prepared as described in the *Immunostaining* section. The One-step TUNEL In Situ Apoptosis Kit (Red, Elab Fluor® 647, E-CK-A324, Elabscience, USA) was used to assess for apoptosis, following the manufacturer’s instructions.

### Glucose-stimulated insulin secretion

Secreted and total insulin content were measured using the Homogeneous Time-Resolved Fluorescence Insulin High Range Detection Kit (62IN1PEG, Revvity, USA) following the manufacturer’s instructions.

### Intracellular Ca^2+^ imaging

Islet-like clusters were incubated with the cytosolic Cal590 dye in KREBS buffer 3.3 mM glucose for 45 minutes at 37°C and 5% CO_2_ before imaging, as described in Supplementary Methods. Images were captured at 1.64 Hz, on a Zeiss AxioObserver Z1 spinning disc system equipped with a Fluor 20X/0.4 DICII at 561 nm, in low glucose (3.3 mM), followed by 11 mM glucose, 16.7 mM glucose and 20 mM KCl.

### Statistical analyses

Integrated multimodal analyses were performed using models including genotype and treatment effects to resolve their respective contributions across omics datasets. Data were analyzed using one-way ANOVA followed by Tukey’s multiple comparison test (GraphPad Prism 10. 6. 1, GraphPad Software, Inc., USA). All data were plotted as mean ± standard deviation (SD).

## Results

### R138X islet-like clusters differentiate normally into insulin producing cells

The R138X mutation was introduced by CRISPR/Cas9 editing into the *SLC30A8* locus of the human embryonic stem cells MEL1, carrying an insulin-GFP reporter, as described [8]. One heterozygous cell line (c.265C>T/c.265C) and one homozygous cell line (c.265C>T/c.265C>T) were generated (**Figure 1a**). Islet-like clusters carrying the *SLC30A8* R138X variant were classified as wild-type (R138X^+/+^), heterozygous (R138X^+/-^), or homozygous (R138X^-/-^). As expected, ZNT8 immunoreactivity was not detected in R138X^-/-^ clusters whereas R138X^+/-^ clusters showed decreased protein levels (**Figure 1b and S1a-b**). After the differentiation and intracellular Zn^2+^ depletion with TPEN, the differentiation efficiency of the newly formed islet-like clusters was evaluated by immunostaining for insulin (INS), NKX6 homeobox 1 (NKX6.1), and glucagon (GCG, **Figure 1b**). Each genotype showed similar numbers of INS^+^ cells (**Figure 1c**). Comparable levels of NKX6.1^+^ cells were detected for R138X^+/+^ and R138X^+/-^ clusters (**Figure 1d**). R138X^-/-^ clusters showed a significant increase of NKX6.1^+^ cells compared to R138X^+/+^, without significant impact on the proportion of INS^+^/NKX6.1^+^ cells (**Figure 1d-e**). Under vehicle conditions, R138X^+/-^ but not R138X^-/-^ clusters showed a slight increase in GCG^+^ cells compared to the R138X^+/+^ clusters (**Figure 1f**). Comparable levels of GCG^+^ and polyhormonal double positive INS^+^/GCG^+^ cells were observed across genotypes (**Figure 1g**). Collectively, these results suggest a subtle Zn^2+^-dependent remodeling of endocrine cell composition.

**Figure 1.**
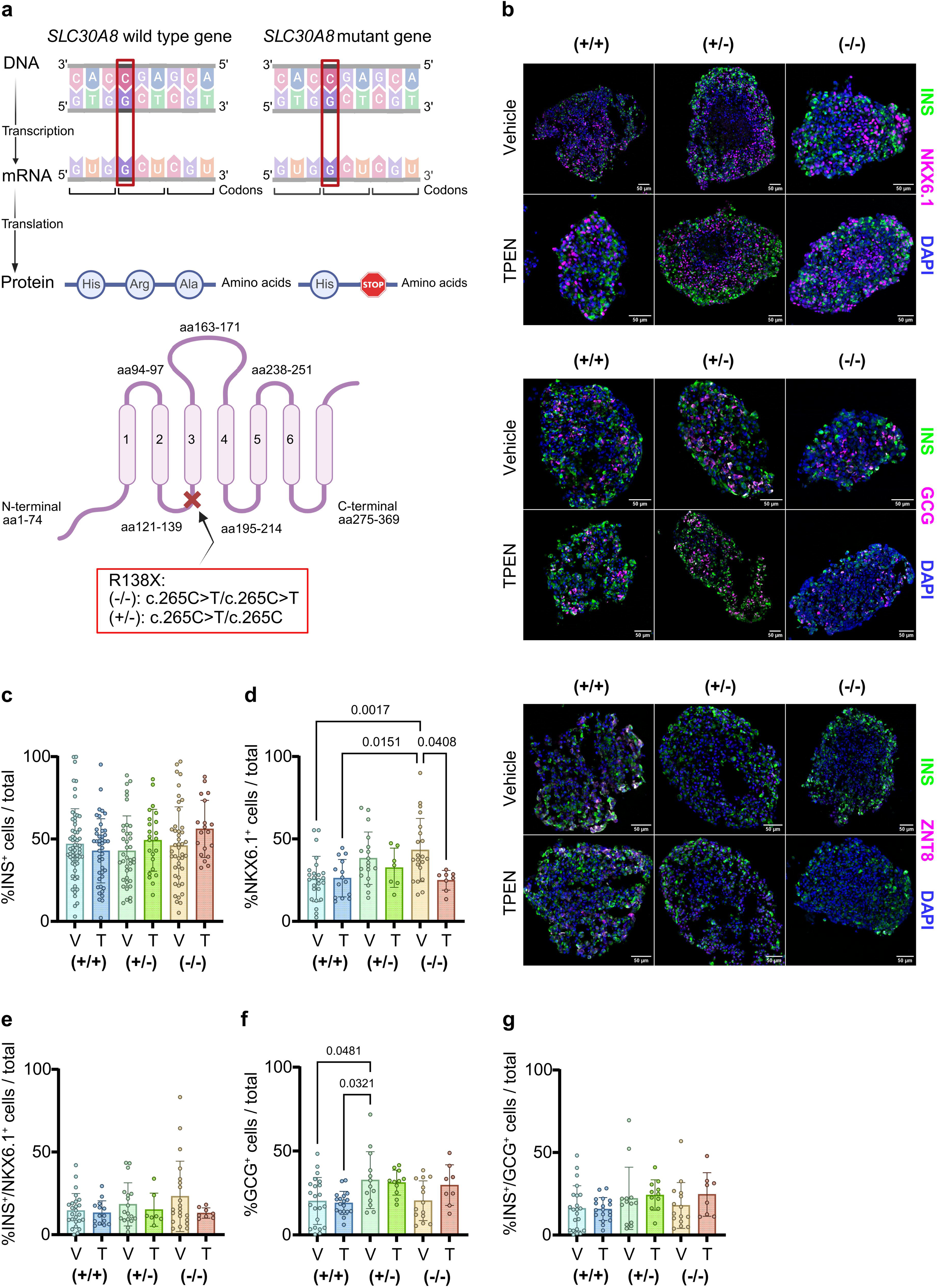
R138X islet-like clusters differentiate normally in insulin producing cells. (**a**) Schematic representation of the *SLC30A8* R138X mutation, leading to the protein truncation, and schematic representation of the SLC30A8 protein with the homozygous (-/-) (c.265C > T/c.265C > T) and heterozygous (+/-) (c.265C > T/c.265C) mutations. (**b**) Representative immunofluorescent images of R138X^+/+^, R138X^+/-^ and R138X^-/-^ islet-like clusters after 27 days of differentiation and 48 hours of TPEN treatment (1 μM), stained for insulin (INS), NKX6 homeobox 1 (NKX6.1), glucagon (GCG), ZNT8 and DAPI. Scale bar = 50 μm. (**c-g**) Immunofluorescent image quantification of INS^+^ cells, NKX6.1^+^ cells, INS^+^/NKX6.1^+^ double positive cells, GCG^+^ cells, and INS^+^/NKX6.1^+^ double positive cells of R138X^+/+^, R138X^+/-^ and R138X^-/-^ islet-like clusters (6 independent differentiations). One-way ANOVA followed by Tukey post hoc test were used to analyze differences groups and data are represented as mean ± SD.

### The absence of active ZnT8 drives proteomic changes in islet-like clusters

To explore how the R138X mutation confers protections against apoptosis in Zn^2+^-depleted conditions, proteomics was performed in islet-like clusters under control or Zn^2+^-depleted conditions. Differential protein expression analysis was conducted using a linear model including both genotype and treatment effects (genotype ∼ treatment), to assess genotype-dependent changes while accounting for Zn^2+^ depletion. A total of 8,971 unique proteins were identified across all samples. Compared with R138X^+/+^ clusters, and accounting for Zn^2+^ depletion, 568 proteins were upregulated (log_2_FC>1, adj. p-value<0.05) and 662 proteins were downregulated (log_2_FC<-1, adj. p-value<0.05) in R138X^+/-^ clusters (**Figure 2a, Table S1**). Similarly, 806 proteins were upregulated (log_2_FC>1, adj. p-value<0.05) and 905 downregulated (log_2_FC<-1, adj. p-value<0.05) in R138X^-/-^ clusters versus R138X^+/+^ clusters (**Figure 2b, Table S2**). Of these, 1,135 proteins were commonly differentially expressed in both mutant genotypes (**Figure S2a**). In contrast, Zn^2+^ depletion (TPEN versus vehicle), assessed while accounting for genotype, resulted in 14 upregulated proteins upregulated (log_2_FC>1) and 17 downregulated (log_2_FC<-1) proteins, but did not reach statistical significance (**Figure S1c-d, Table S3**).

**Figure 2.**
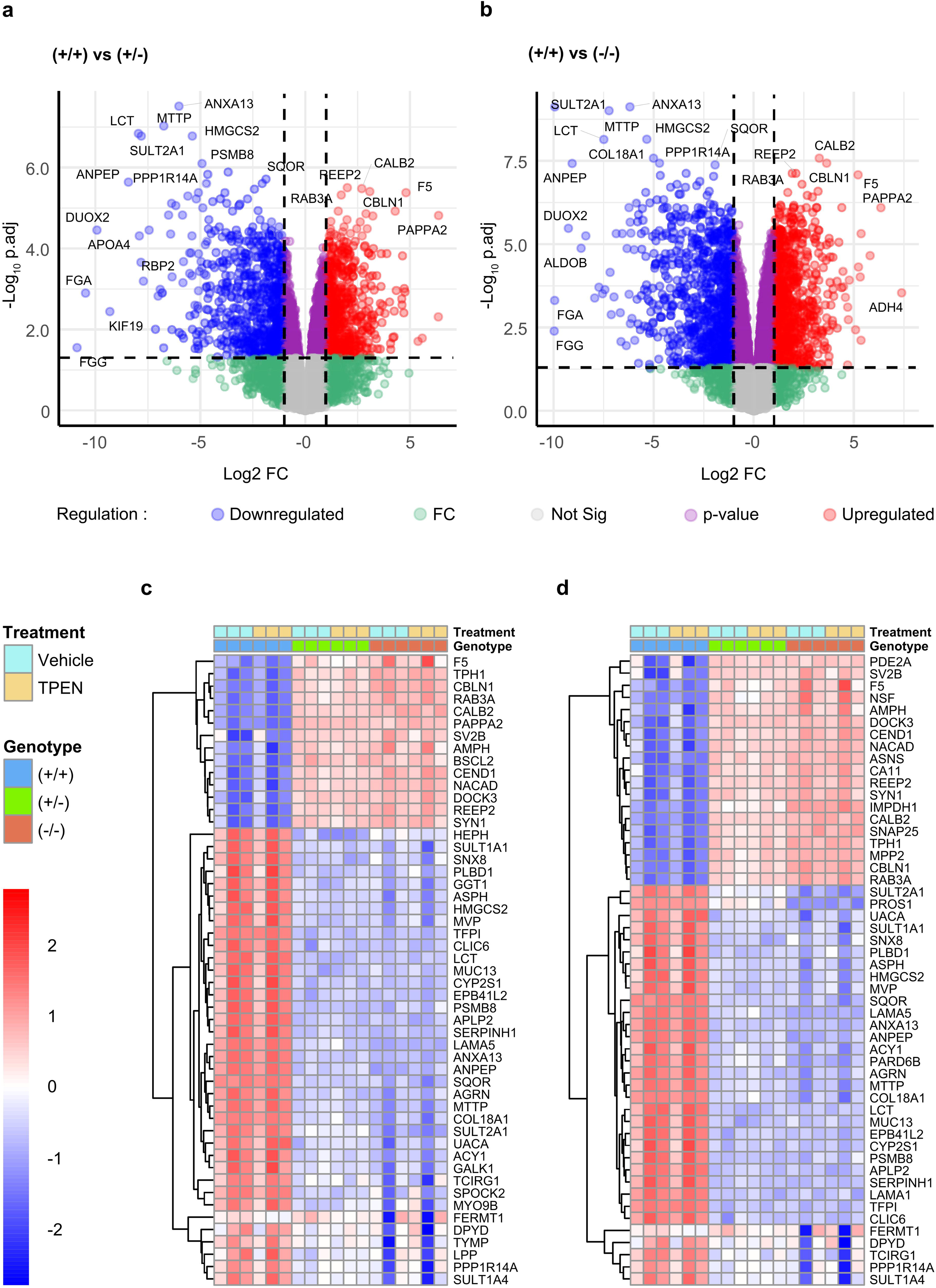
The absence of active ZnT8 drives proteomic changes in R138X islet-like clusters. (**a-b**) Volcano plots showing differential protein expression between R138X^+/+^ and R138X^+/-^ clusters (**a**) and between R138X^+/+^ and R138X^-/-^ clusters (**b**), while accounting zinc depletion treatment (TPEN 1 μM, n = 3 independent differentiations). (**c-d**) Hierarchical clustering representing the top 50 most significantly altered proteins between R138X^+/+^ and R138X^+/-^ clusters (**c**) and between R138X^+/+^ and R138X^-/-^clusters (**d**), as revealed by limma analysis. Data were clustered by Ward’s clustering with the Euclidean distance in R. Heatmap columns represent each genotype, in vehicle or TPEN conditions, and rows represent dysregulated proteins.

Clustering analysis revealed groups of dysregulated proteins in each genotype. Among the 50 top dysregulated proteins in the R138X^+/-^ clusters, proteins involved in vesicle trafficking and insulin exocytosis, including RAB3A, SYN1, SV2B and AMPH, and in intracellular Ca^2+^ homeostasis, such as CALB2 and ASPH, were upregulated (**Figure 2c**). Interestingly, the protein responsible for IGF binding protein cleavage, the metalloprotease PAPPA2, was strongly upregulated, suggesting a possible increase in local IGF1 levels. In parallel, proteins involved in the maintenance of the extracellular matrix (ECM) and cellular structure (LAMA5, COL18A1, AGRN and FERMT1), stress and chaperone proteins (SERPINH1, PSMB8, UACA and HEPH) and metabolic enzymes (HMGCS2 (ketogenesis), SULT1A1/2A1/1A4 (detoxification)) were downregulated. These effects were exacerbated in R138X^-/-^ clusters, with

SNAP25, NSF, DOCK3 and CBLN1 completing the list of upregulated proteins associated with vesicle trafficking and exocytosis and highlighted with the heterozygous genotype (**Figure 2d**). Identical observations were made regarding the downregulated proteins related to ECM, cellular stress, and metabolic proteins, complemented by proteins associated with cellular transport and membranes (MTTP, LCT, CLIC6, PLBD1, EPB41L2). While both heterozygous and homozygous R138X mutations led to significant proteomic alterations, the magnitude of these changes was greater in homozygous clusters, supporting a dose-dependent effect of *SLC30A8*.

To support these results, Gene Set Enrichment Analyses (GSEA) were performed and the top dysregulated pathways are represented in **Figure 3**. In both the heterozygous and homozygous phenotypes, the most upregulated pathways were associated with *vesicle trafficking and secretion*, and *energy metabolism* (**Figures 3a-3c, Tables S4-S5**). The most downregulated pathways included pathways related to ECM involving collagen-related proteins (COL6A1, COL6A2, COL2A1, COL4A2), matrix metallopeptidases (MMPs; MMP14, MMP11, MMP2, MMP15) and laminins (LAMA5, LAMA1, LAMA3). *Metabolism* pathways were also disrupted and involved apolipoproteins (APOC1, APOA4, APOB, APOA2, APOE) and proteoglycans (SDC1, SDC2, SDC4, GPC3, HSPG2) (**Figures 3b-3d, Tables S4-S5**). Altogether, our data highlight a strong proteome remodeling, suggesting a reprogramming of cellular functions in response to *SLC30A8* mutations, with a shift towards enhanced secretory and bioenergetic activity at the expense of structural and homeostatic pathways.

**Figure 3.**
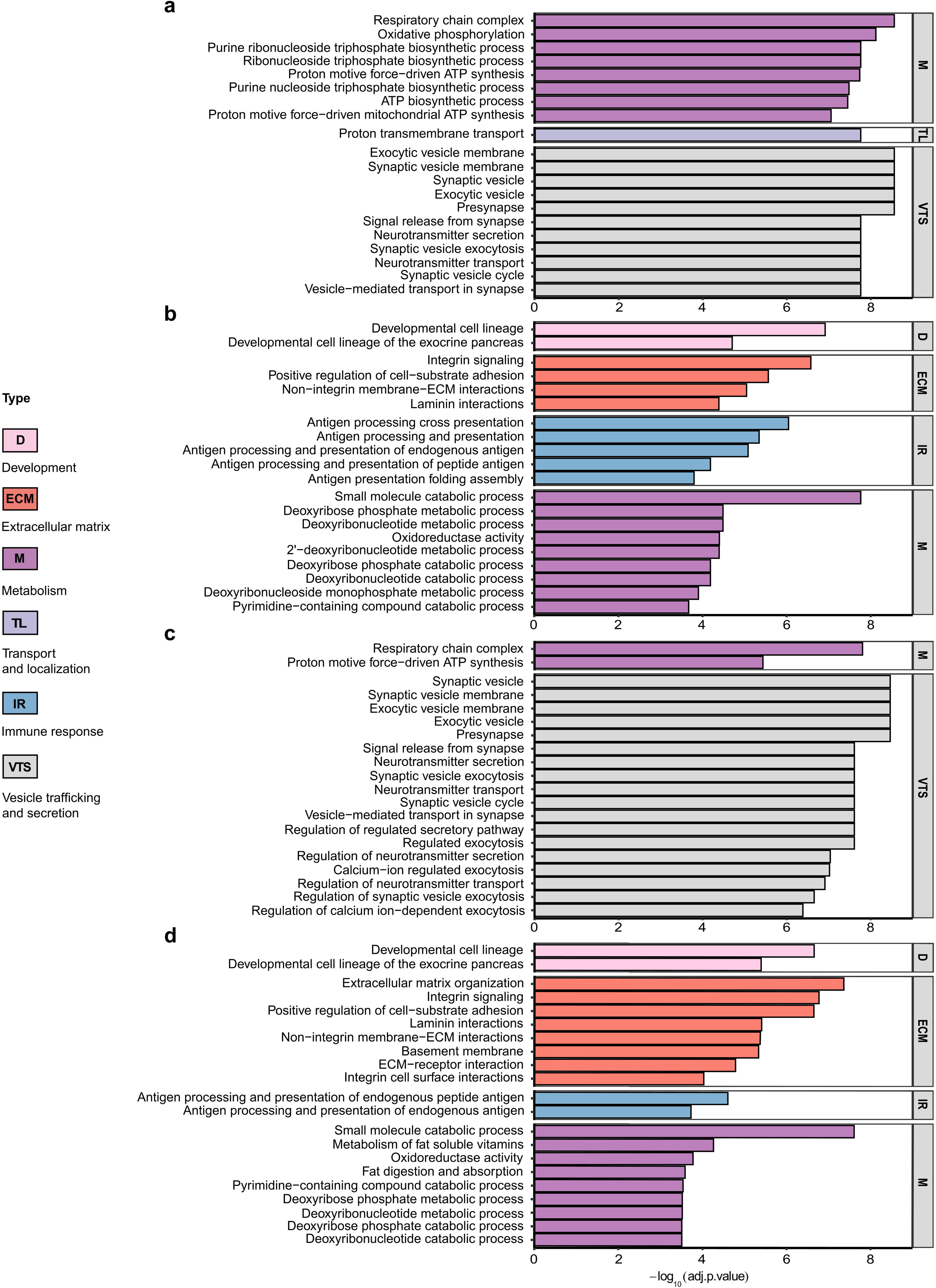
The proteomic changes altered major signaling pathways in R138X islet-like clusters. Top 20 most upregulated (**a** and **c**) and downregulated (**b** and **d**) signaling pathways between R138X^+/+^ and R138X^+/-^ clusters (**a-b**) and between R138X^+/+^ and R138X^-/-^ clusters (**c-d**), as revealed by Gene Set Enrichment Analysis. The top dysregulated pathways were selected according to the *Normalized Enrichment Score* and an adjusted q-value < 0.05. For improved biological relevance, pathways containing more than 500 proteins were removed to avoid overly broad and non-specific categories.

### Proteomic changes reflect alterations in the transcriptome

To further decipher the impact of functional ZnT8 in islet-like clusters, we performed bulk mRNA sequencing in fully differentiated clusters after Zn^2+^-depletion. Consistent with proteomic analysis, differential gene expression analysis was conducted using a linear model including both genotype and treatment effects (genotype ∼ treatment). A total of 61,951 genes were identified in all samples. Compared with R138X^+/+^ clusters, and accounting for Zn^2+^ depletion, 929 genes were upregulated (log_2_FC>1, adj. p-value<0.05) and 1,557 genes downregulated (log_2_FC<-1, adj. p-value<0.05) in R138X^+/-^ clusters (**Figure 4a, Table S6**). Similarly, 1,418 genes were upregulated (log_2_FC>1, adj. p-value<0.05) and 2,396 downregulated (log_2_FC<-1, adj. p-value<0.05) in R138X^-/-^ clusters versus R138X^+/+^ clusters (**Figure 4b, Table S7**). Of these, 1,926 genes were commonly differentially expressed in both mutant genotypes (**Figure S2b**). Cross-omics integration identified 264 and 493 overlapping genes between proteomic and transcriptomic datasets in R138X^+/-^ and R138X^-/-^ clusters, respectively (**Figure S2c-d**). Finally, Zn^2+^ depletion (TPEN versus vehicle), assessed while accounting for genotype, resulted in 1,162 upregulated (log_2_FC>1) and 1,284 downregulated (log_2_FC<-1) genes, but did not reach statistical significance (**Figure S1e-f, Table S8**).

**Figure 4.**
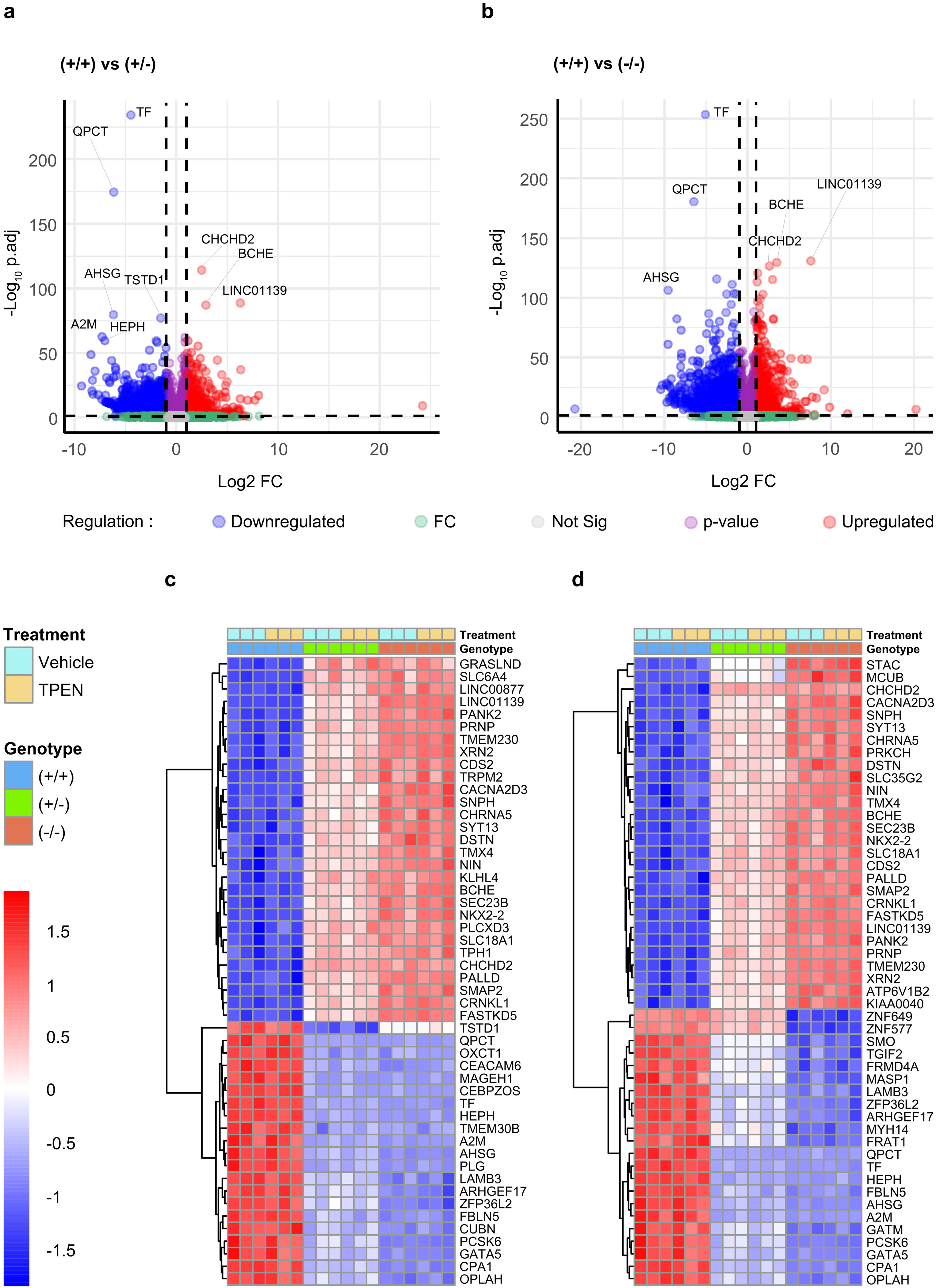
Transcriptomic analyses support an alteration of ion channels, exocytosis and ECM in R138X clusters. (**a-b**) Volcano plots showing differential gene expressions between R138X^+/+^ and R138X^+/-^ clusters (**a**) and between R138X^+/+^ and R138X^-/-^clusters (**b**), while accounting zinc depletion (TPEN 1 μM, n = 3 independent differentiations). (**c-d**) Hierarchical clustering representing the top 50 most significantly altered genes between R138X^+/+^ and R138X^+/-^ clusters (**c**) and between R138X^+/+^ and R138X^-/-^ clusters (**d**), as revealed by DESeq analysis. Data were clustered by Ward’s clustering with the Euclidean distance in R. Heatmap columns represent each genotype, in vehicle or TPEN conditions, and rows represent dysregulated proteins.

Both R138X^+/-^ and R138X^-/-^ clusters showed largely similar sets of genes among the top 50 most dysregulated. Clustering analysis revealed a coordinated upregulation of genes associated with calcium-dependent exocytosis (*SYT13*, *SLC18A1*, *CACNA2D3*, *SMAP2*, *SEC23B*), mitochondrial respiration and energy metabolism (*PANK2*, *CHCHD2*, *FASTKD5*, *CDS2*) and beta cell identity (*NKX2.2*) (**Figures 4c-d**). Alongside, genes associated with extracellular matrix components (*LAMB3*, *FBLN5*), oxidative stress and metabolism (*HEPH*, *TF*, *OPLAH*), transcription regulation (*GATA5*, *ZFP36L2*) and exocrine pancreas markers (*CPA1*) were downregulated. In line with the proteomic results, GSEA revealed that for both genotypes, top upregulated signaling pathways were associated with *vesicle trafficking and signaling*, *cell signaling and stress*, *gene expression*, and *transport and localization* (**Figure 5a-5c, Tables S9-S10**). The top downregulated signaling pathways were associated with *extracellular matrix*, *cell signaling* and *development* (**Figure 5b-5d, Tables S9-S10**). Collectively, the omics data support a shift toward a more functionally mature, metabolically active, and endocrine-specialized islet cell state, accompanied by remodeling of the islet microenvironment.

**Figure 5.**
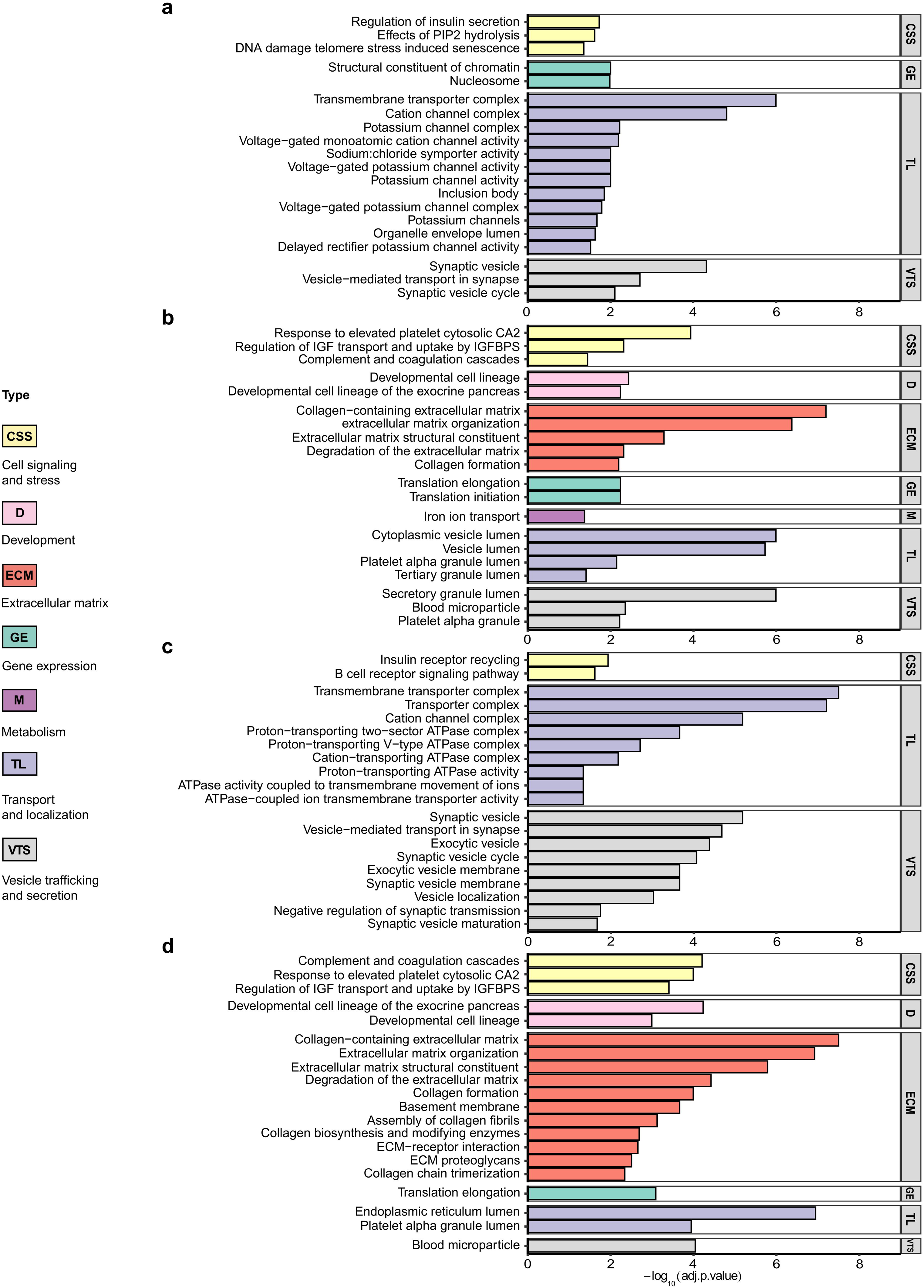
Transcriptomic changes lead to the remodeling of major signaling pathways in R138X islet-like clusters. Top 20 most upregulated (**a** and **c**) and downregulated (**b** and **d**) signaling pathways between R138X^+/+^ and R138X^+/-^ clusters (**a-b**) and between R138X^+/+^ and R138X^-/-^ clusters (**c-d**), as revealed by Gene Set Enrichment Analysis. The top dysregulated pathways were selected according to the *Normalized Enrichment Score* and an adjusted q-value < 0.05. For improved biological relevance, pathways containing more than 500 proteins were removed to avoid overly broad and non-specific categories.

### Effects of ZnT8 deficiency on beta cell and endocrine markers

To gain further understanding of the impact of ZnT8 deficiency, we next focused on genes associated with beta cell identity and endocrine markers. Full differential expression results from omic analysis are provided in **Table S11**. Several genes associated with beta cell identity and function, including *NKX6.1*, *NEUROD1*, *MNX1*, *IGF1R*, *IAPP*, *GLP1R* and *FOXA2* were upregulated in R138X^+/-^ and R138X^-/-^ clusters (**Figure 6a**). *NPY* and *ALDH1A3*, both markers of failing beta cells, were downregulated in the mutant clusters, supporting a more mature state in the absence of ZnT8. In line with this, several genes associated with endocrine lineage, including *UCN3*, *PYY*, *PPY*, *PAX6*, *PAX4*, *NKX2.2* and *GCG*, were upregulated (**Figure 6b**). Genes associated with the pancreatic progenitor stage were downregulated, including *SOX9*, *NEUROG3*, *GATA4* and *CPA1* (**Figure 6c**). Interestingly, several disallowed genes (**Figure S3a**) and proteins (**Figure S3b**) were downregulated in the mutant clusters, including LDHA and SLC16A1, supporting a reduced conversion of glucose to lactate and a shift toward oxidative metabolism, as highlighted by GSEA analyses (**Figures 3 and 5**).

**Figure 6.**
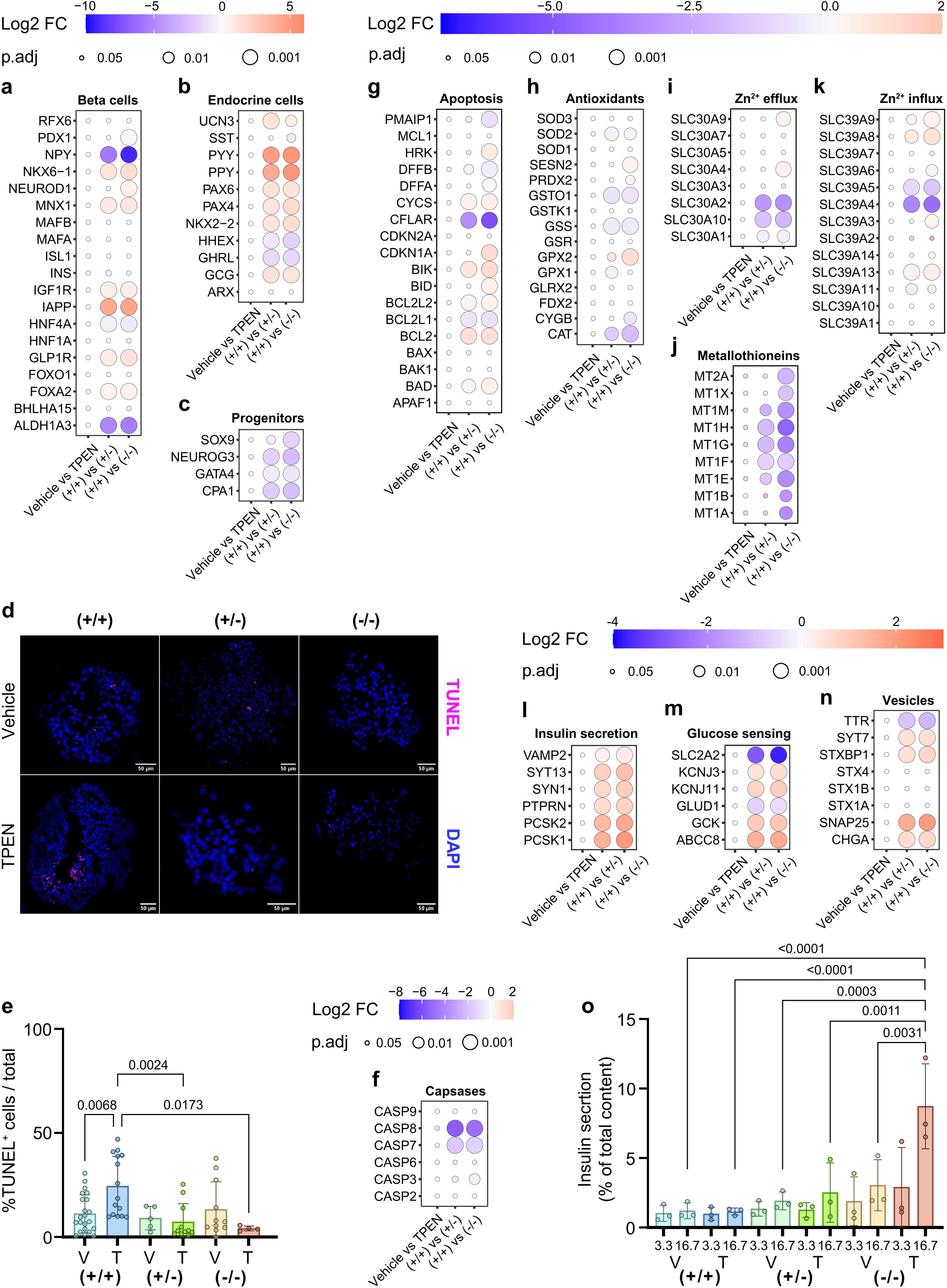
The absence of ZnT8 disrupted genes and proteins associated with various islet functions without inducing apoptosis. (**a-c**) Expression levels of key beta cell (**a**), endocrine (**b**), and pancreatic progenitor (**c**) genes. The dot colors represent the log_2_FC and the dot size represents the adjusted p-value, as revealed by the DESeq analysis of the transcriptomic data. The treatment effect was evaluated while accounting the genotype (Vehicle vs TPEN). The genotype effect was evaluated while accounting zinc depletion, and comparison were made between R138X^+/+^ and R138X^+/-^ clusters ((+/+) vs (+/-)) and between R138X^+/+^ and R138X^-/-^ clusters ((+/+) vs (-/-)). (**d**) Representative immunofluorescent images of R138X^+/+^, R138X^+/-^ and R138X^-/-^ islet-like clusters after 27 days of differentiation and 48 hours of TPEN treatment (1 μM), stained for TUNEL and DAPI. Scale bar = 50 μm. (**e**) Immunofluorescent image quantification of TUNEL^+^ cells of R138X^+/+^, R138X^+/-^ and R138X^-/-^ islet-like clusters (from 6 differentiations). One-way ANOVA followed by Tukey post hoc test were used to analyze differences groups and data are represented as mean ± SD. (**f**) Protein expression levels of caspases, as revealed by limma analysis of the proteomic data. Similar to transcriptomic results, the treatment effect was evaluated while accounting the genotype (Vehicle vs TPEN). The genotype effect was evaluated while accounting zinc depletion, and comparison were made between R138X^+/+^ and R138X^+/-^ clusters ((+/+) vs (+/-)) and between R138X^+/+^ and R138X^-/-^clusters ((+/+) vs (-/-)). (**g-k**) Expression levels of key apoptotic (**g**), antioxidant (**h**), and zinc efflux (**i**), metallothioneins (**j**) and zinc influx (**k**) genes, as revealed by the DESeq analysis of the transcriptomic data. (**l-n**) Expression levels of genes involved in insulin secretion (**l**), glucose sensing (**m**) and vesicle exocytosis (**n**). (**o**) Percentage of secreted insulin relative to total insulin content from R138X^+/+^, R138X^+/-^ and R138X^-/-^ islet-like clusters under basal glucose (3.3 mM) and high glucose (16.7 mM) after 27 days of differentiation and 48 hours of TPEN treatment (1 μM, n = 3 independent differentiations). Two-way ANOVA was used to analyze differences groups and data are represented as mean ± SD.

### R138X clusters are protected against apoptosis and cellular stress

The primary function of beta cell to secrete insulin is closely related to Zn^2+^ transport, and zinc plays a key role in the regulation of apoptosis. In Zn^2+^-depleted conditions, R138X^+/+^ clusters showed increased apoptosis, as demonstrated by TUNEL staining (p=0.0068, **Figure 6d-e**). However, R138X^+/-^ and R138X^-/-^ clusters were protected against this induction (**Figure 6d**). At the protein level, this was associated with a significant decrease of CASP7 and CASP8, as revealed by proteomics (**Figure 6f**). At the transcriptomic level, the anti-apoptotic gene *BCL2* was increased while the apoptosis regulator *CFLAR* was decreased (**Figure 6g**). Importantly, expression of endoplasmic reticulum (ER) stress-related genes, including *PDIA6*, *HYOU1*, *CRYAB*, *ATF3* and *XBP1*, were downregulated by the absence of ZnT8, suggesting a lowered ER stress and unfolded protein response (**Figure S3c**). Several antioxidant genes, such as *CAT*, *GPX2*, *GSS* and *GSTO1*, and metallothioneins were also strongly downregulated, suggesting an adaptive response to preserve cell survival under reduced Zn^2+^ availability (**Figure 6h-6j**). Metallothioneins are also involved in Zn^2+^ transport, and their alteration was associated with dysregulation of *Solute Carrier Family 30* and *Family 39 genes*, involved in Zn^2+^ efflux and influx, indicating altered Zn^2+^ handling in islet-like clusters lacking ZnT8 (**Figure 6i-6k**). ZNT8 protein levels were decreased in the R138X^+/-^ clusters and not detected in the R138X^-/-^ clusters, as revealed by the proteomic analysis (**Table S11**).

### R138X clusters have increased insulin secretion

In addition to these effects, genes associated with insulin secretion (*PCSK1*, *PCSK2*, *SYN* and *SYT13*), glucose sensing (*GCK*, *KCNJ11* and *ABCC8*), vesicle exocytosis (*CHGA* and *SNAP25*) and the paracrine peptide *EDN3* were all upregulated in R138X mutant clusters (**Figure 6l-n and S3d**). This transcriptional profile was accompanied by lowered expression of the glucose transporter *SLC2A2* and several hormone-related genes, including *PRSS23*, *APOE* and *APOA1* (**Figure 6m and S3d**). Measurements of insulin secretion corroborated these findings, showing a significant increase in insulin release from R138X^-/-^ clusters under Zn^2+^-depleted, glucose-stimulated conditions, compared to the other cell line responses, regardless of TPEN treatment (**Figure 6o and S3e**).

### Zinc depletion alters Ca^2+^ signaling and cell-to-cell connectivity in islet-like clusters

To validate our findings above, cytosolic Ca^2+^ signaling was measured in islet-like clusters in vehicle and TPEN conditions at cellular resolution (**Figure 7a**). After glucose stimulation at 11 mM (11G) or 16.7 mM (16.7G), cytosolic Ca^2+^ revealed an expected increase in R138X^+/+^ clusters (**Figure 7b-c**). In TPEN conditions, R138X^+/+^ clusters showed a decreased responsiveness to high glucose, as demonstrated by a decrease in the incremental area under the curve (AUC) at 11G (p=0.0168) and amplitude of the oscillations at 16.7G (p=0.0012) at the cellular level (**Figure 7h and m**). R138X^+/-^ and R138X^-/-^ clusters demonstrated altered Ca^2+^ dynamics compared to R138X^+/+^ clusters (**Figure 7d-g**), with a minor decrease in AUC at 11G (p<0.0001) and 16.7G (p=0.0041 and 0.0007, respectively for each genotype) and a significant increase of oscillation frequencies at 11G (p<0.0001) and 16.7G (p=0.001 and 0.0006, respectively) in vehicle conditions (**Figure 7h-k**). However, R138X^-/-^ clusters also showed lowered amplitude at 11G (p=0.0002) and 16.7G (p=0.0020) compared to the R138X^+/+^ clusters. Interestingly, TPEN treatment reversed these effects on AUC, especially at 16.7G (**Figure 7h-i**). In R138X^+/-^ clusters, the frequency of the oscillations was even further increased (p<0.0001 vs R138X^+/+^+T clusters; **Figure 7j-k**) and an amplitude similar to the R138X^+/+^ clusters in vehicle condition was observed (p=0.0418 at 11G; p<0.0001 at 16.7G; vs R138X^+/+^+T clusters; **Figure 7l-m**). In R138X^-/-^ clusters, the frequency and amplitude of the oscillations were similar to those in R138X^+/+^ clusters in vehicle conditions. No changes in peak width were observed in any conditions (**Figure S4a-b**).

**Figure 7.**
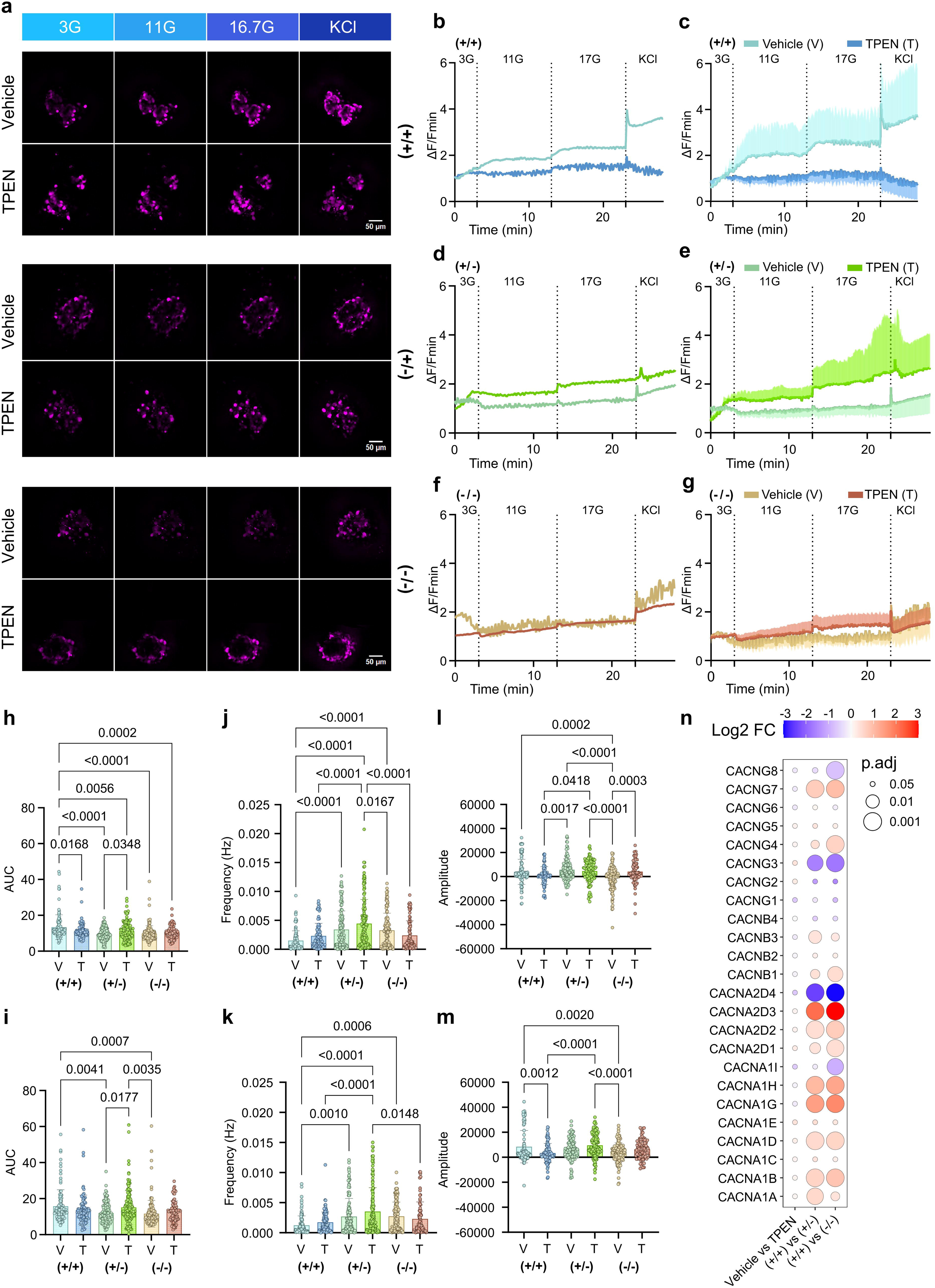
Zinc depletion alters intracellular calcium signaling in islet-like clusters. (**a**) Snapshots from cytosolic Ca^2+^ imaging of R138X^+/+^, R138X^+/-^ and R138X^-/-^ islet-like clusters after 27 days of differentiation and 48 hours of TPEN treatment (1 μM), at the indicated glucose concentrations. Scale bar = 50 μm. (**b-g**) Corresponding cytosolic Ca^2+^ traces from the whole islets (**b**, **d**, and **f**) and from individual cells within each islet (**c**, **e** and **g**) in response to 3.3 mM glucose (3G), 11 mM glucose (11G), 16.7 mM glucose (16.7G) and 20 mM KCl in R138X^+/+^ (**b-c**), R138X^+/-^ (**d-e**) and R138X^-/-^ islet-like clusters (**f-g**). (**h-i**) Corresponding Ca^2+^ area under the curve (AUC) at 11G (**h**) and 16.7G (**i**) from all individual cells across all analyzed islets. (**j-k**) Corresponding Ca^2+^ oscillation frequencies at 11G (**j**) and 16.7G (**k**) from all individual cells across all analyzed islets. (**l-m**) Corresponding Ca^2+^ oscillation amplitude at 11G (**l**) and 16.7G (**m**) from all individual cells across all analyzed islets. Calcium dynamics were assessed using Cal-590 for cytosolic Ca²⁺. Traces represent mean normalized fluorescence intensity over time (ΔF/F_min_). For AUC, frequency and amplitude analyses, measurements were performed between 3–13 min at 11G and 13–23 min at 16.7G. One-way ANOVA followed by Tukey post hoc test were used to analyze differences groups and data are represented as mean ± SD. (**n**) Expression levels of voltage-gated calcium channel genes. The dot colors represent the log_2_FC and the dot size represents the adjusted p-value, as revealed by the DESeq analysis of the transcriptomic data. The treatment effect was evaluated while accounting the genotype (Vehicle vs TPEN). The genotype effect was evaluated while accounting zinc depletion, and comparison were made between R138X^+/+^ and R138X^+/-^ clusters ((+/+) vs (+/-)) and between R138X^+/+^ and R138X^-/-^ clusters ((+/+) vs (-/-)).

The aforementioned observations were made at the cellular level, without major changes at the islet level (**Figure S4c-f**). Consistent with these changes in Ca^2+^ dynamics, the expression profile of voltage-gated calcium channel subunits revealed a selective upregulation of major components involved in Ca^2+^ influx, including *CACNA1D/1G/1H* and regulatory subunits such as *CACNA2D1/D2/D3* (**Figure 7n**). Moreover, genes involved in mitochondrial Ca^2+^ handling, including *MCUB*, *UCP2*, *SMFT1*, and *MICU3* were upregulated, suggesting a potential remodeling of mitochondrial Ca^2+^ dynamics (**Figure S4i**). In contrast, mitochondrial life cycle genes, including *PRKN*, *PPARGC1B*, *PPARD*, *PPARA* and *LITAF*, were downregulated, consistent with altered mitochondrial turnover (**Figure S4k**). Finally, several genes involved in ER Ca^2+^ handling, including *STIM1*, *STIM2*, *RYR2* and *ITPR1/2/3*, were also increased (**Figure S4j**). Altogether, these transcriptional changes reflected a coordinated adaptation of ER and mitochondrial Ca^2+^ handling.

Given these alterations, we next explored cell-cell connectivity in islet-like clusters using intracellular cytosolic Ca^2+^ dynamics as a proxy for cellular activity [24]. Raster plots of ΔF/F_min_ revealed heterogenous Ca^2+^ activity during glucose stimulation, regardless of glucose concentration (**Figure 8a**). Comparing with R138X^+/+^ clusters under vehicle conditions, both R138X^+/-^ and R138X^-/-^ clusters exhibited lowered overall Ca^2+^ activity. Despite this lowered activity, R138X^-/-^ clusters showed increased functional connectivity already at 11G (**Figure 8b**). Upon Zn^2+^ depletion, activity was reduced in R138X^+/+^ clusters. In contrast, TPEN increased Ca^2+^ activity, apparent connectivity (**Figure 8a–b**), and Pearson’s correlation coefficient in R138X^+/-^ clusters (**Figure 8c**), without affecting cellular coactivity (**Figure S4g–h**). In parallel, under vehicle conditions, R138X^-/-^ clusters displayed a significant increase in duty cycle and delayed time to response compared to R138X^+/+^ clusters (**Figure 8c**). Overall, these results indicate that ZnT8 deficiency in R138X mutant clusters alters Ca^2+^ dynamics, and that Zn^2+^ depletion partially restores these functional changes.

**Figure 8.**
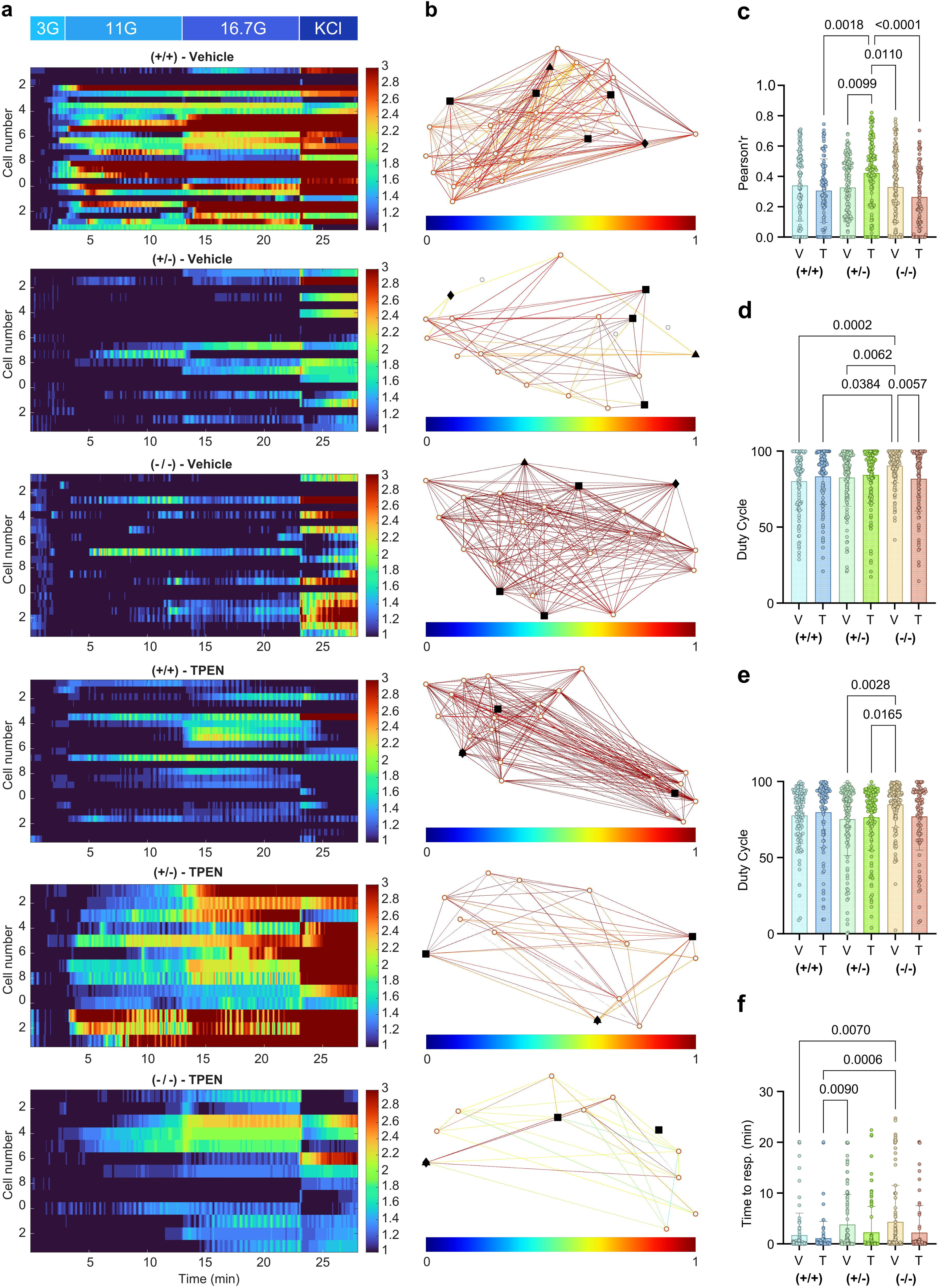
ZnT8 absence and zinc depletion modulate cell-cell connectivity in R138X islets. (**a**) Raster plots corresponding to the cytosolic Ca^2+^ ΔF/F_min_ fluorescent intensity over time for individual cells from R138X^+/+^, R138X^+/-^ and R138X^-/-^ islet-like clusters after 27 days of differentiation and 48 hours of TPEN treatment (1 μM), at the indicated glucose concentrations. Corresponding movies are provided in supplementary materials. (**b**) Representative Cartesian map of beta cells from islets in (**a**) with color-coded lines connecting cells according to the strength of connectivity at 11G (color-coded values from 0 to 1, blue to red). Beta cells represented with the triangle are the first responders; cells represented with the diamond shape are showing an earlier onset of intracellular Ca^2+^ elevation; the square represents the best-connected cells. (**c**) Pearson’r correlation associated with the synchronicity of the oscillations. (**d-e**) Duty cycle of the Ca^2+^ oscillations at 11G (**d**) and 16.7G (**e**). (**f**) Time to respond (in minutes) for each cell. Analyses were performed in individual islet-like clusters (n = 3 individual differentiations). Data are presented as mean ± SD. One-way ANOVA followed by Tukey post hoc test were used to analyze differences groups and data are represented as mean ± SD.

## Discussion

The present study aimed to dissect the molecular mechanisms by which an inactivating, truncating variant in *SLC30A8* (p.Arg138*, R138X) dose-dependently protects against type 2 diabetes [15]. To this end, we deploy an unbiased proteomic approach, complementing earlier transcriptomic analyses [8], in CRISPR-edited human stem cell-derived islet-like clusters carrying heterozygous or homozygous R138X mutations. We further assessed whether altered susceptibility to Zn^2+^ depletion-induced dysfunction or apoptosis is involved in the action of this variant [14].

Both transcriptomic and proteomic changes were strongly dose-dependent, consistent with diabetes risk [15]. In line with previous studies, ZnT8 loss did not impair normal differentiation, with or without Zn^2+^ depletion [8, 18], but instead promoted a shift towards greater maturity, as supported by the increase of genes associated with beta cell function and identity and other endocrine markers, and decreased pancreatic progenitor markers. We observed a decrease in *neuropeptide Y* (*NPY*) and *Aldehyde dehydrogenase 1 Family member A3* (*ALDH1A3*) expression, both markers of immature beta cells, as well as several disallowed genes [25, 26]. Conversely, we observed a strong increase in *Endothelin 3* (*EDN3*) expression, recently identified as a key regulator of beta cell identity and insulin expression under hypoxia [27]. These changes were associated with increased expression of other genes involved in beta cell maturation and glucose sensing, and with enhanced glucose-induced insulin secretion (GSIS) (**Figure 6**), despite reduced *SLC2A2* (*GLUT2*) expression and activation of energy metabolism and exocytosis pathways (**Figure 3**), particularly under Zn^2+^-depleted conditions.

Zn^2+^ depletion alone itself had limited effects at either the transcriptomic or proteomic level (**Figure S1**), suggesting that its primary impact may occur through post-translational mechanisms. However, it markedly modulated the effects of the inactivating ZnT8 mutation. Notably, Zn^2+^ depletion revealed pronounced effects of the R138X variant on both apoptosis (**Figure 6b-c**) and on GSIS (**Figure 6g**), which were subtle or absent under normal conditions. These findings are reminiscent of a relationship between Zn^2+^ intake and the effects of the common R325W variant on diabetes risk [11] and suggest that susceptibility to dietary zinc deficiency may be mitigated in carrier of inactivating ZnT8 mutations.

Functionaly, R138X clusters displayed lowered Ca^2+^ activity but preserved insulin secretion, consistent with our previous findings [8]. Interestingly, under TPEN-induced Zn^2+^ depletion, Ca^2+^ responses, connectivity and insulin secretion were enhanced (**Figures 6-8**). These adaptations were associated with the upregulation of key components of voltage-gated calcium channels (*CACNA1D*, *CACNA1G*, *CACNA1H*, *CACNA2D1-3*), ER Ca^2+^ regulators (*STIM1*, *STIM2*, *RYR2*, *ITPR1/2/3*) and mitochondrial Ca^2+^ handling genes (*MCUB*, *UCP2*, *SMDT1*, *MICU3*). Interestingly, ER Gla protein (ERGP), encoded by ASPH [28], was strongly downregulated in R138X^+/-^and R138X^-/-^ clusters, suggesting a role for altered ER Ca2+ homeostasis in the effects of the variant. Together, these data suggest a rewiring of intracellular Ca^2+^ dynamics, allowing R138X clusters to maintain their Ca^2+^ signaling despite altered Zn^2+^ homeostasis.

We further demonstrated a marked protective effect of the ZnT8 R138X variant against apoptosis induced by Zn^2+^ depletion, in agreement with previous studies [8, 17, 29]. This was associated with an overall downregulation of zinc handling pathways, including zinc transporters of the SLC family and metallothioneins, and decreased signatures of oxidative and ER stress (**Figures 6e and S3c**). Importantly, previous studies have demonstrated that ZnT8 loss alters overall Zn^2+^ distribution rather than cytosolic Zn^2+^ levels. Indeed, ZnT8 deficiency leads to reduced granular Zn^2+^ content and crystallized insulin granules, without major changes in cytoplasmic Zn^2+^ content [8]. *In vivo*, this was accompanied by a redistribution of Zn^2+^ at the tissue level in R138X mice, with decreased endocrine Zn^2+^ content and increased exocrine accumulation, resulting in a more homogenous pancreatic Zn^2+^ distribution [19].

Proteomic analyses further identified several factors contributing to enhanced cell survival. These included Pappalysin 2 (PAPPA2), which increases IGF availability through IGF-binding protein cleavage, thereby promoting pro-survival signaling pathways [30], and anti-apoptotic proteins such as BCL2. Notably, approximatively 50% of ZnT8 protein has been reported to localize to the ER, and it has been shown to protect against cytokine-induced cytotoxicity and glucolipotoxicity by attenuating ER stress [18, 29]. Consistent with this, we observed lowered ER stress-related pathways, suggesting that ZnT8 deficiency could attenuate key stress responses involved in beta cell failure.

Finally, our data point to a previously unidentified role of ZnT8 in regulating tissue architecture. ZnT8 depletion was associated with extensive remodeling of the ECM and cytoskeletal networks. Given that Zn^2+^ is an important cofactor for matrix metalloproteinases (MMPs), which mediate the degradation of ECM proteins, reduced intracellular Zn^2+^ availability may enhance ECM turnover and remodeling [31]. Our detailed proteomic analysis revealed alterations in multiple pathways linked to tissue organization and cytoskeletal dynamics. These included component of the Hedgehog signaling pathway (SMO), several regulators of actin cytoskeleton architecture (DSTN, PALLADIN, ARHGEF17, FRDM4, MYH14), collagen-related proteins (COL6A1, COL6A2, COL2A1, COL4A2), matrix metallopeptidases (MMPs: MMP14, MMP11, MMP2, MMP15) and laminins (LAMA5, LAMA1, LAMA3). Together, these changes suggest a coordinated structural remodeling which may contribute to enhanced beta cell survival in the absence of ZnT8 in Zn^2+^ depleted conditions.

We note that our findings are in line with earlier results showing that mice bearing the R138X mutation show normal body weight, glucose tolerance and beta cell mass, but a 50% increase in insulin secretion in hyperglycemic conditions. Taken together with results in humans [14, 15], and the present findings in stem cell-derived islet-like clusters, these results show that the absence of ZnT8 may exert beneficial effects of glucose metabolism in diabetic-like conditions [32].

### Limitations

Our transcriptomic analyses were performed using bulk mRNA sequencing, which may mask cell-type specific effects. However, in our previous study [8], we performed single-cell RNA sequencing in R138X islet-like clusters, in the absence of Zn^2+^-depleted conditions, and our new results align and complement our previous observations. In the omic analyses, TPEN treatment, hence Zn^2+^ depletion had little effect on gene and protein levels. Given the strong impact of R138X mutation on islet-like clusters, the treatment effect might be masked by the genotype effect. However, functional analyses were able to reveal the effect of Zn^2+^ depletion on insulin secretion, Ca^2+^ signaling and cell-cell connectivity, and the induction of apoptosis, indicating that acute Zn^2+^ chelation mostly leads to post-translational modifications. Future studies using chronic Zn^2+^ modulation throughout differentiation will be required.

## Conclusion

Our results support a model in which ZnT8 loss-of-function induces various cellular and metabolic adaptations, integrating possible Zn^2+^ redistribution, Ca^2+^ signaling modulation, stress and apoptosis lowering and structural remodeling in human islet-like clusters. Rather than lowering intracellular Zn^2+^ levels, Zn^2+^ depletion or the absence of ZnT8 reshape both intracellular and extracellular microenvironments. Altogether, these findings highlight the therapeutic potential of targeting ZnT8 in type 2 diabetes, and of ensuring appropriate dietary zinc intake.

## Data availability

Bulk mRNA sequencing data will be deposited in the NCBI Sequence Read Archive (SRA). The mass spectrometry proteomics data will be deposited at the ProteomeXchange Consortium via the PRIDE (DOI: 10.1093/nar/gkab1038).

## Acknowledgments

Cellular imaging was performed using the Microscopy platform of the CRCHUM (Centre de Recherche du Centre Hospitalier de l’Université de Montréal) and we gratefully acknowledge Aurélie Cleret-Buhot (Plateforme d’imagerie cellulaire, CRCHUM) for her expert assistance. Proteomic analysis was performed by the Mass Spectrometry and Proteomics Platform (Montreal Clinical Research Institute, IRCM) and we thank Denis Faubert and Josée Champagne for their help and guidance throughout the project. Proteomic data bioinformatics analyses were performed at the Bioinformatics core facility from IRCM and we thank Virginie Calderon for her expertise. We thank Dax Fu for providing the mAb20 antibody for ZnT8 immunostaining. We also gratefully thank Alexandre Haldemann for his help and support to design the figures.

## Contributions

MG co-designed the study, performed all the experiments and biostatistics, with the exception of mRNA sequencing and LC-MS/MS proteomics and proteomic data analyses, wrote, reviewed and edited the manuscript, and generated all the figures. IC provided training and support for stem-cell differentiation, immunocytochemistry and insulin secretion experiments. GO contributed to calcium imaging and connectivity analyses. MF contributed to proteomic data generation and analyses. QD and DE provided the human embryonic stem cell lines, MEL1 and CRISPR/Cas9 edited lines. GAR co-designed the study, supervised the project, reviewed and edited the manuscript and provided funding. All authors carefully reviewed the manuscript and gave their approval regarding the final version. GAR serves as the guarantor of this study.

## Abbreviations

Ca^2+^: calcium ion
ECM: extracellular matrix
ER: endoplasmic reticulum
GSEA: gene set enrichment analysis
SLC30A8: Solute Carrier Family 30 member 8 (ZnT8)
SLC39A8: Solute Carrier Family 39 member 8
TPEN: N,N,N′,N′-tetrakis(2-pyridylmethyl)-1,2-ethanediamine
V: vehicle
Zn^2+^: zinc ion

---

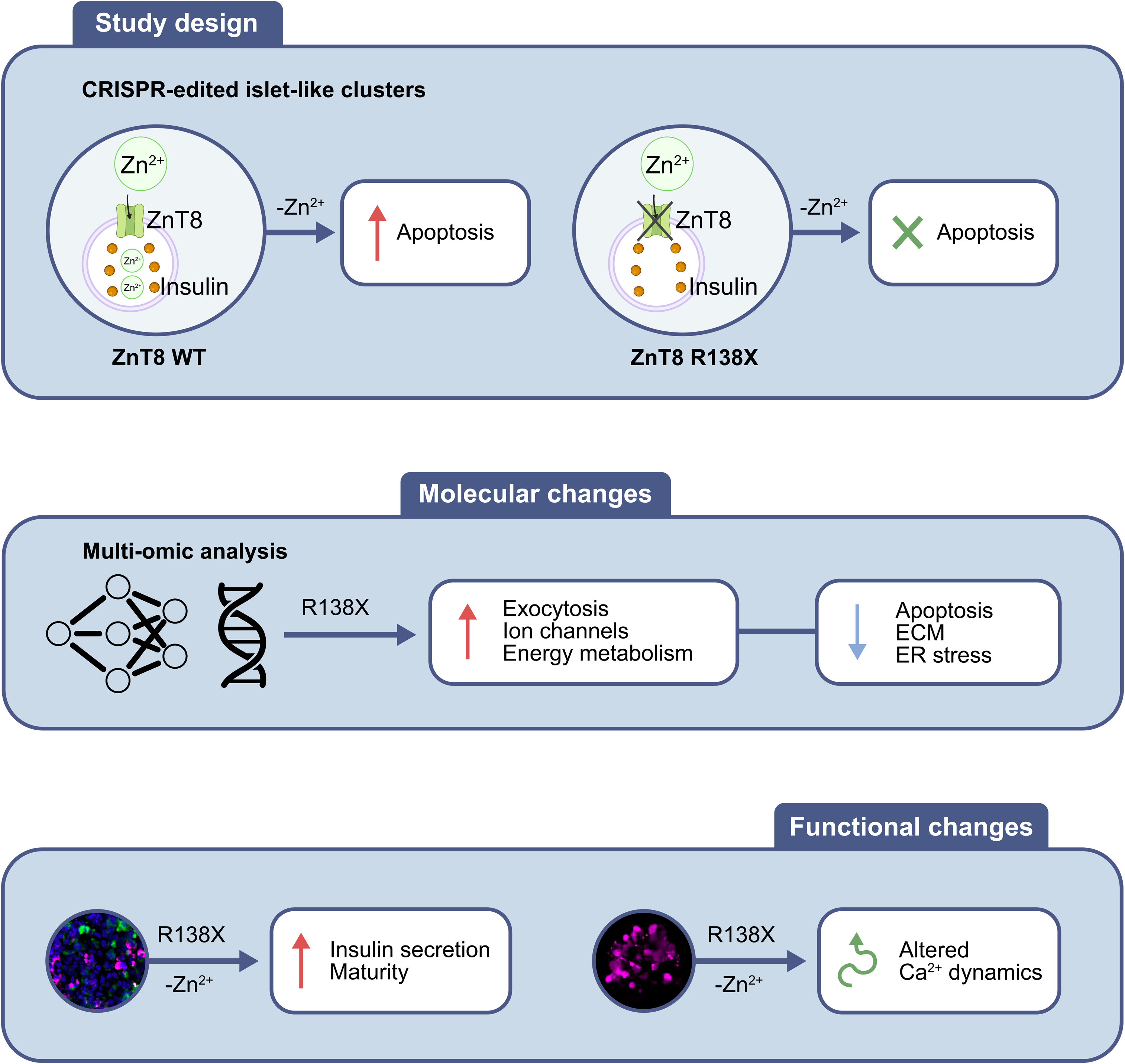

