## Supplementary information for "A loss of function variant in *SLC30A8/ZnT8* drives proteomic changes associated with lowered apoptosis in human stem cell-derived islets"

### Extended methods

#### ***Differentiation of human embryonic stem cells***

Human embryonic stem cells were obtained as described [1]. Cell lines with mutations were established by introducing heterozygous and homozygous p.Arg138* mutations into human embryonic stem cell line MEL1 (NIH registry #0139) with using CRISPR/Cas9 [1]. The common SLC30A8 variant rs13266634 (R325W) has been associated with beta cell dysfunction and type 2 diabetes risk. However, the genotype of the MEL1 human embryonic stem cell line at this locus is not reported in the literature and was not determined in the present study.

Human embryonic stem cells were expanded in complete StemFlex medium (Cat. No. A334901, Thermo Fisher Scientific, USA) at 37°C and 5% $CO_2$. Cells were seeded into 6-well plates at 2.1 x $10^5$ cells/$cm^2$ in complete StemFlex medium as described [2]. At 100% confluence, definitive endoderm stage was induced and cells were cultured for 4 days using the STEMdiff™ Definitive Endoderm Differentiation Kit (Cat. No. 05110, STEMCELL Technologies, Canada). The primitive gut tube stage was induced from day 4 to day 6 by RPMI 1640 plus GlutaMAX™ (Cat. No. 61870127, Thermo Fisher Scientific) supplemented with 1% (v/v) Penicillin- Streptomycin (P/S, Cat. No. 15140122, Thermo Fisher Scientific), 1% (v/v) B-27 serum-free supplement (50X, Cat. No. 17504044, Thermo Fisher Scientific) and 50 ng/mL Human FGF-7 (KGF) Recombinant Protein (KGF, Cat. No. PHG0094, Thermo Fisher Scientific). The posterior foregut stage was induced from day 6 to day 8 by DMEM, high glucose, GlutaMax™ Supplement (Cat. No. 10566024, Thermo Fisher Scientific), supplemented with 1% (v/v) P/S, 1% (v/v) B-27, 50 ng/mL KGF, 0.25 μM Cyclopamine-KAAD (Cat. No. 239804-100UG, Sigma-Aldrich, USA), 2 μM Retinoic acid (Cat. No.

72262, STEMCELL Technologies) and 0.25 µM LDN193189 (Cat. No. SML0559, Sigma-Aldrich). From day 8 to day 12, we generated pancreatic progenitor cells with DMEM plus GlutaMax, 1% (v/v) P/S, 1% (v/v) B-27, 25 ng/mL KGF and 50 ng/mL Epidermal Growth Factor (EGF, Cat. No. 236-EG-200, R&D Systems Inc, USA).

On day 12, cells were dissociated into single cells using TrypLE™ Express (Cat. No. 12605036, Life Technologies) and clustered in AggreWell 400 6-well plates (Cat. No. 34425, STEMCELL Technologies) with DMEM plus GlutaMax supplemented with 1% (v/v) P/S), 1% (v/v) B-27, 10 ng/mL KGF, 1 µM ALK5 inhibitor (Repsox, Cat. No. 73794, STEMCELL Technologies) and 10 µM Y-27632 (ROCK inhibitor, Cat. No. M1817-10MG, AbMole BioSciences, USA). On day 13, newly formed clusters were transferred into ultra-low attachment 6-well plates (Cat. No. 07-200-601, Thermo Fisher Scientific) in RPMI 1640 plus GlutaMAX™ supplemented with 1% (v/v) P/S, 1% (v/v) B-27, 1 µM thyroid hormone (T3) (Cat. No. T6397, Sigma-Aldrich), 10 µM Repsox, 10 µM zinc sulfate (Cat. No. AAJ60963AE, Thermo Fisher Scientific), 10 µg/mL heparin (Cat. No. H3149-10KU, Sigma-Aldrich), 100 nM gamma-secretase inhibitor (Cat. No. 565789-500UG, Sigma-Aldrich), 10 µM ROCK inhibitor for 7 days. From day 15, 1 µM aphidicolin (Cat No. A4487, Sigma-Aldrich) was added to the medium for differentiation improvement. Finally, to induce pancreatic beta cell, medium was changed to RPMI 1640 plus GlutaMAX™ supplemented with 1% (v/v) P/S, 1% (v/v) B-27, 10% (v/v) Fetal Bovine Serum (FBS, Cat No. 098-150, Canada) and 1 µM aphidicolin from day 20 to day 27.

***Immunostaining***

After 48 hours of TPEN treatment, clusters were fixed with 4% paraformaldehyde (PFA) at room temperature (RT) for 15 minutes. Clusters were washed with Dulbecco's phosphate-buffered saline, no calcium, no magnesium (DPBS, Cat. No. 14190144, Thermo Fisher Scientific) and incubated overnight in 30% sucrose (Cat. No. S0389, Sigma-Aldrich). Clusters were then transferred into a cryomold and frozen in O.C.T. compound (Cat. No. 23-730-571, Thermo Fisher Scientific) at -80°C. 10 µm sections were cut from the frozen blocks on microscope slides using a cryostat. Slides were stored at -80°C until staining.

On staining day, clusters were circled with a hydrophobic pen and slides were rehydrated in Phosphate Buffer Saline (PBS, Cat. No. P4417, Sigma-Aldrich) for 15 minutes. Membranes were permeabilized with cold methanol for 10 minutes at -20°C. Slides were washed 3 times with PBS and blocked in 2% Bovine Serum Albumin (BSA, Cat. No. A7030, Sigma-Aldrich) in PBS-Tween (Cat. No. P1379, Sigma-Aldrich) 0.1% for 1 hour at RT, covered with parafilm in a humidified chamber. Slides were incubated with primary antibodies (**Table S12**) in 2% BSA in PBS-Tween 0.1% overnight at 4°C. Slides were then washed 3 times with PBS-Tween 0.1% and incubated with secondary antibodies (**Table S13**) in 2% BSA in PBS-Tween 0.1% for 45 minutes at RT. Slides were then washed 3 times with PBS-Tween 0.1% and finally incubated with DAPI (**Table S13**) in 2% BSA in PBS-Tween 0.1% for 10 minutes at RT. Slides were mounted and stored at 4°C before imaging. Pictures were taken with a confocal Leica Stellaris 8 microscope with a 20X/0.75 objective. Data were analysed using QuPath software [3]. Due to limited sample size (n=1–2 per condition) for ZNT8 staining, no statistical analysis was performed and data were presented descriptively.

***Proteomics***

*Protein extraction*

Islet-like clusters were washed twice with ice cold PBS using gentle centrifugations. Proteins were extracted on ice with 50 µL of Lysis Buffer (20 mM Tris-HCl pH 7.5 (Cat. No. 15506017, Thermo Fisher Scientific), 150 mM NaCl (Cat. No. SOD001.205, BioShop, Canada), 1 mM EDTA (Cat. No. E5134, Sigma-Aldrich) pH 8.0, 1 mM EGTA (Cat. No. E3889, Sigma-Aldrich) pH 8.0, 2.5 mM NaPyrophosphate (Cas. No. A17546.30, Thermo Fisher Scientific), 1 mM beta-glycerophosphate (Cat. No. 35675, Sigma-Aldrich), 10 mM NaF (Cat. No. 201154, Sigma-Aldrich), 1% Triton (Cat. No. X100, Sigma-Aldrich), 1 mM phenylmethylsulfonyl fluoride protease inhibitor (PMSF, Cat. No. 36978, Thermo Fisher Scientific), 1X protease inhibitor (Cat. No. 78430, Thermo Fisher Scientific); pH 7.5; sterilized with a 0.45 µm filter). Proteins were incubated on ice for 10 minutes and store in lysis buffer at -80°C until further analyses. protein concentrations were quantified with the Pierce BCA Protein Assay Kit following manufacturer instructions. 20 µg of proteins were used for protein extraction and LC-MS/MS injection.

*On-Bead Digestion using Protein Aggregation Capture (PAC) beads*

Samples were initially reduced with 9 mM dithiothreitol (DTT) in 50 mM Triethylammonium bicarbonate buffer (TEAB) incubated on an Eppendorf ThermoMixer at 37°C for 30 minutes at 700 RPM. After cooling for 5 minutes, samples were alkylated with 17 mM iodoacetamide (IAA) in 50 mM TEAB at room temperature for 30 minutes at 700 RPM, protected from light. To remove residual nucleic acids, samples were treated with 50 units of benzonase per sample in the presence of 2 mM $MgCl_2$, incubated at 37°C for 60 minutes at 700 RPM. Protein Aggregation Capture

(PAC) was performed using 7,5 µL of washed Hydroxyl beads (ReSyn Biosciences) per sample. 100% acetonitrile was added to achieve a final concentration of 50%, initiating protein aggregation. Samples were incubated at room temperature for 20 minutes at 1000 RPM, then placed on a magnetic rack for 2 minutes. Supernatants were discarded and retained for further analysis. Beads were washed three times with 70% ethanol. Following the final wash, beads were resuspended in 100 µL of 0.01 µg/µL Lys-C/Trypsin mix (Promega) in 50 mM TEAB. Samples were briefly sonicated in a water bath for 1 minute and digested overnight at 37°C, shaking at 700 RPM. Upon completion of digestion, samples were placed on a magnetic rack, and the supernatants were transferred to Eppendorf LoBind tubes. Beads were rinsed with 25 µL of 50 mM TEAB, and rinsed supernatants were pooled with the initial digests. They were acidified at 0.5% formic acid, dried with a Speed-Vac and kept at -20°C until ready for the LC-MS/MS analysis.

*LC-MS/MS Analysis*

Dried peptides were reconstituted in 30 µL of 2% acetonitrile and 1% formic acid, agitated for 15 minutes, and 450 ng of each sample was injected for analysis. Peptides were separated on a 75 µm inner diameter × 170 mm self-packed C18 column (Dr. Maisch) installed on a Vanquish Neo liquid chromatography system (Thermo Scientific). Mobile phases consisted of 0.2% formic acid in water (buffer A) and 85% acetonitrile with 0.2% formic acid (buffer B). Peptides were eluted at a flow rate of 250 nL/min using a two-step linear gradient of 3–50% buffer B over 60 minutes, followed by 50–90% buffer B over 4 minutes.

The LC system was coupled to an Orbitrap Astral mass spectrometer (Thermo Scientific) via a Nanospray Flex Ion Source. Spray voltage was set to 1.3–1.5 kV, the RF lens to 40 V, and the ion transfer tube temperature to 250 °C. Full MS scans were acquired in the Orbitrap over an m/z range of 380–980 at a resolution of 240,000, with a target intensity of $5 \times 10^6$ and a maximum injection time of 3 ms. Data-independent acquisition (DIA) scans were acquired in the Astral analyzer over the same m/z range, using a target intensity of $5 \times 10^4$ and a maximum injection time of 3 ms. DIA isolation windows were set to 2 Th, and fragmentation was performed using a normalized collision energy of 25%.

*Protein Identification and Quantification*

Raw data files were processed in Proteome Discoverer v3.2.0.450 (Thermo Scientific) using a CHIMERYS-based DIA workflow. Spectra were selected using the Spectrum Selector node with precursor reevaluation based on isotope patterns enabled, a minimum peak count of five, and an FT-MS signal-to-noise threshold of 1.5.

Peptide identification was performed using CHIMERYS on an Ardia server, searching against the UniProt Homo sapiens Swiss-Prot canonical protein database (TaxID 9606, release 404), supplemented with common contaminants. Peptides were generated using trypsin with full specificity, allowing up to one missed cleavage. The minimum and maximum peptide lengths were set to 7 and 30 amino acids, respectively, with allowed precursor charge states from +2 to +4. Carbamidomethylation of cysteine residues was specified as a fixed modification, while oxidation of methionine was included as a variable modification, with a maximum of three variable modifications per peptide. Fragment ion mass tolerance was set to 20 ppm. Peptide-spectrum matches (PSMs) were validated using a target–decoy approach, with strict and relaxed false discovery rate (FDR) thresholds set to 1% and 5%, respectively.

Following identification, results were consolidated in a consensus workflow including PSM grouping, peptide validation, protein filtering, and strict parsimony-based protein grouping. Only high-confidence peptides and master proteins were retained. Protein-level FDR was controlled at 1% (strict). Protein annotation included Gene Ontology biological process, molecular function, and cellular component terms.

Quantification was performed using the Fragment Ions Quantifier node, using unique and razor peptides, with precursor intensities derived from MS2 apex intensities. Protein abundances were calculated as the sum of peptide abundances, excluding modified peptides from ratio calculations. No normalization or imputation was applied during quantification. Final results were filtered to retain high-confidence master proteins for downstream analysis.

*Statistical analysis*

The output from Proteome Discoverer was provided by the IRCM proteomics platform. 8,971 unique proteins were identified in all samples. First, intensities were normalized using Variance Stabilizing Normalization (VSN). Data were then filtered to retain proteins detected in at least 50% of the samples and quantified with at least 2 peptides giving 8,971 proteins. Differential analysis was carried out using limma R package on normalized intensities, without imputation of missing values to identify DAPs (Differentially Abundant Proteins). Missing values were imputed with the 1st intensity percentile of each protein (noise value) only for visualization to allow data representation (PCA, heatmap, volcano). Functional enrichment analysis of DAPs was performed using clusterProfiler with Reactome, KEGG, and Gene Ontology databases. Genes were ranked based on $log_2$ fold change and adjusted p-values, and pathways were classified as up- or downregulated based on normalized enrichment scores (NES). Analyses were performed with R-4.5.2. Bioinformatics analyses were performed at the Bioinformatics core facility from Montreal Clinical Research Institute (IRCM).

***RNA-Sequencing***

Total mRNA was isolated from using the RNeasy Mini Kit (Cat. No. 74104, Qiagen, Netherlands) following the manufacturer's protocol. RNA concentrations were quantified using the Nanodrop (Thermo Fisher Scientific, USA). 500 ng of total RNA were used as input for bulk mRNA Sequencing.

RNA sequencing was performed by Genome Québec (Montreal, Canada). Transcript quantification was performed using Salmon and imported into R with tximport, summarizing counts at the gene level using Ensembl GRCh38 v115 annotations. Quality control was conducted on raw counts, including total read counts, number of detected genes, and mitochondrial (MT-) and ribosomal (RPL/RPS) gene proportions. Housekeeping gene expression was assessed across samples. Batch effects were corrected using ComBat-seq. Principal component analysis was performed on log-transformed counts to evaluate sample clustering. Differential gene expression analysis was carried out using DESeq2 with models including genotype and treatment effects. Counts were normalized using the median-of-ratios method, and statistical significance was determined using the Wald test with Benjamini–Hochberg correction (FDR < 0.05). Gene set enrichment analysis (GSEA) was performed using clusterProfiler with Reactome, KEGG, and Gene Ontology databases. Genes were ranked based on $log_2$ fold change and adjusted p-values, and pathways were classified as up- or downregulated based on normalized enrichment scores (NES). Analyses were performed with R-4.5.2.

***TUNEL assay***

Pictures were taken with a confocal Leica Stellaris 8 microscope with a 20X/0.75 objective. Data were analyses using QuPath software.

***Glucose-stimulated insulin secretion***

15 clusters of similar size were selected per cell line, +/- treatment, and transferred into 1.5 mL Eppendorf tubes. Clusters were pre-incubated for 1 hour in KREBS buffer (Krebs-Ringer bicarbonate (KREBS) buffer (140 mM NaCl, 3.6 mM KCl, 0.5 mM $NaH_2PO_4$, 24 mM $NaHCO_3$, 1.5 mM $CaCl_2$, 0.5 mM $MgSO_4$, 10 mM HEPES and 3 mM D-glucose; pH 7.4; 1% BSA, pH 7.4), 3.3 mM glucose, sterilized with a 0.22 µm filter, at 37°C and 5% $CO_2$. The supernatant was removed and clusters were again incubated for 30 minutes in KREBS buffer, 3.3 mM glucose. Supernatant was collected and directly stored at -80°C. Clusters were washed once with KREBS buffer no glucose and incubated for 30 minutes in KREBS buffer, 16.7 mM glucose. Supernatant was collected and directly stored at -80°C. Clusters were washed once with KREBS buffer no glucose and incubated for 30 minutes in KREBS buffer, 3.3 mM glucose, 20 mM KCl. Supernatant was collected and directly stored at -80°C. 50 µL of $ddH_2O$ were added to the remaining clusters. Clusters were then sonicated for 1 second at 20% power, and total insulin content was extracted by adding 150 µL of acid EtOH and incubated overnight at 4°C. Clusters were vortexed and centrifuged for 10 minutes at 4°C and 10,000g. Total insulin content were stored at -80°C until further analyses. Secreted and total insulin content were measured using the HTRF Insulin High Range Detection Kit (62IN1PEG, Revvity, USA) following manufacturer instructions.

#### *Intracellular $Ca^{2+}$ imaging*

Data were analysed using ImageJ v1. 54p [4] and MatLab v25.2.0.3042426 (Natick, USA; 2022.). Calcium traces of insulin were analyzed for connectivity as previously described [5], with optimization for islet-like clusters: a signal threshold of $\Delta F/F>2\%$ was used.

#### *Statistical analyses*

The differences observed were evaluated as statistically significant when p-value was less than 0.05 and were displayed on figures.

#### *References*


[1] Sui L, Du Q, Romer A, et al. (2023) ZnT8 Loss of Function Mutation Increases Resistance of Human Embryonic Stem Cell-Derived Beta Cells to Apoptosis in Low Zinc Condition. Cells 12(6). 10.3390/cells12060903

[2] Cherkaoui I, Du Q, Egli D, Misra S, Rutter GA (2024) Optimized Protocol for Generating Functional Pancreatic Insulin-secreting Cells from Human Pluripotent Stem Cells. J Vis Exp(204). 10.3791/65530

[3] Bankhead P, Loughrey MB, Fernandez JA, et al. (2017) QuPath: Open source software for digital pathology image analysis. Sci Rep 7(1): 16878. 10.1038/s41598-017-17204-5

[4] Schindelin J, Arganda-Carreras I, Frise E, et al. (2012) Fiji: an open-source platform for biological-image analysis. Nat Methods 9(7): 676–682. 10.1038/nmeth.2019

[5] Cherkaoui IG, M.; Du, Q.; Egli, D. M.; Dion, C.; Leitch, H. G.; Ostinelli, G.; Sachedina, D.; Misra, S.; Rutter, G. A. (2026) A recessive HNF1A p.A251T variant

causes monogenic diabetes by altering islet cell development, insulin secretion and intercellular connectivity. medRxiv. https://doi.org/10.1101/2024.12.10.24318788

### Supplementary tables

**Table S1.** Dysregulated proteins between R138X$^{+/+}$ and R138X$^{+/-}$ clusters, while taking into account zinc depletion treatment. Differential analysis was carried out using limma R package on normalized intensities.

**Table S2.** Dysregulated proteins between R138X$^{+/+}$ and R138X$^{-/-}$ clusters while, taking into account zinc depletion treatment. Differential analysis was carried out using limma R package on normalized intensities.

**Table S3.** Dysregulated proteins between vehicle and TPEN treatment, while taking into account the cluster genotypes. Differential analysis was carried out using limma R package on normalized intensities.

**Table S4.** Gene Set Enrichment Analysis results between R138X$^{+/+}$ and R138X$^{+/-}$ clusters while, taking into account zinc depletion treatment. Enrichment analysis was performed using clusterProfiler with Reactome, KEGG, and Gene Ontology databases. Proteins were ranked based on $log_2$ fold change and adjusted p-values, and pathways were classified as up- or downregulated based on normalized enrichment scores (NES). For improved biological relevance, pathways containing more than 500 proteins were removed to avoid overly broad and non-specific categories.

**Table S5.** Gene Set Enrichment Analysis results between R138X$^{+/+}$ and R138X$^{-/-}$ clusters while, taking into account zinc depletion treatment. Enrichment analysis was performed using clusterProfiler with Reactome, KEGG, and Gene Ontology databases. Proteins were ranked based on $log_2$ fold change and adjusted p-values, and pathways were classified as up- or downregulated based on normalized enrichment scores (NES). For improved biological relevance, pathways containing more than 500 proteins were removed to avoid overly broad and non-specific categories.

**Table S6.** Dysregulated genes between R138X$^{+/+}$ and R138X$^{+/-}$ clusters, while taking into account zinc depletion treatment. Differential analysis was carried out using DESeq R package on normalized intensities.

**Table S7.** Dysregulated genes between R138X$^{+/+}$ and R138X$^{-/-}$ clusters while, taking into account zinc depletion treatment. Differential analysis was carried out using DESeq R package on normalized intensities.

**Table S8.** Dysregulated genes between vehicle and TPEN treatment, while taking into account the cluster genotypes. Differential analysis was carried out using DESeq R package on normalized intensities.

**Table S9.** Gene Set Enrichment Analysis results between R138X$^{+/+}$ and R138X$^{+/-}$ clusters while, taking into account zinc depletion treatment. Enrichment analysis was performed using clusterProfiler with Reactome, KEGG, and Gene Ontology databases. Genes were ranked based on $\log_2$ fold change and adjusted p-values, and pathways were classified as up- or downregulated based on normalized enrichment scores (NES). For improved biological relevance, pathways containing more than 500 proteins were removed to avoid overly broad and non-specific categories.

**Table S10.** Gene Set Enrichment Analysis results between R138X$^{+/+}$ and R138X$^{-/-}$ clusters while, taking into account zinc depletion treatment. Enrichment analysis was performed using clusterProfiler with Reactome, KEGG, and Gene Ontology databases. Genes were ranked based on $\log_2$ fold change and adjusted p-values, and pathways were classified as up- or downregulated based on normalized enrichment scores (NES). For improved biological relevance, pathways containing more than 500 proteins were removed to avoid overly broad and non-specific categories.

**Table S11.** Full differential expression results for specific gene and protein classes, extracted from the transcriptomic and proteomic data.

**Table S12.** Primary antibody list

| **Primary Antibody List** | | | | |
|---|---|---|---|---|
| Antibody | Species | Dilution | Company | Catalog No. |
| Insulin | Guinea Pig | 1:10 | Agilent | IR00261-2 |
| NKX6.1 | Mouse | 1:50 | DSHB | F55A12 |
| Glucagon | Mouse | 1:1,000 | Sigma-Aldrich | G2654 |
| ZnT8 | Mouse | 20 µg/mL | Dax Fu lab | na |

**Table S13.** Secondary antibody list

| **Secondary Antibody List** | | | |
|---|---|---|---|
| Antibody | Dilution | Company | Catalog No. |
| Alexa Fluor@ 546 Goat anti-guinea pig IgG (H+L) | 1:1,000 | Invitrogen | A11074 |
| Alexa Fluor@ 647 Goat anti-mouse IgG (H+L) | 1:1,000 | Invitrogen | A21235 |
| DAPI | 1:1,000 | Sigma-Aldrich | D9542 |

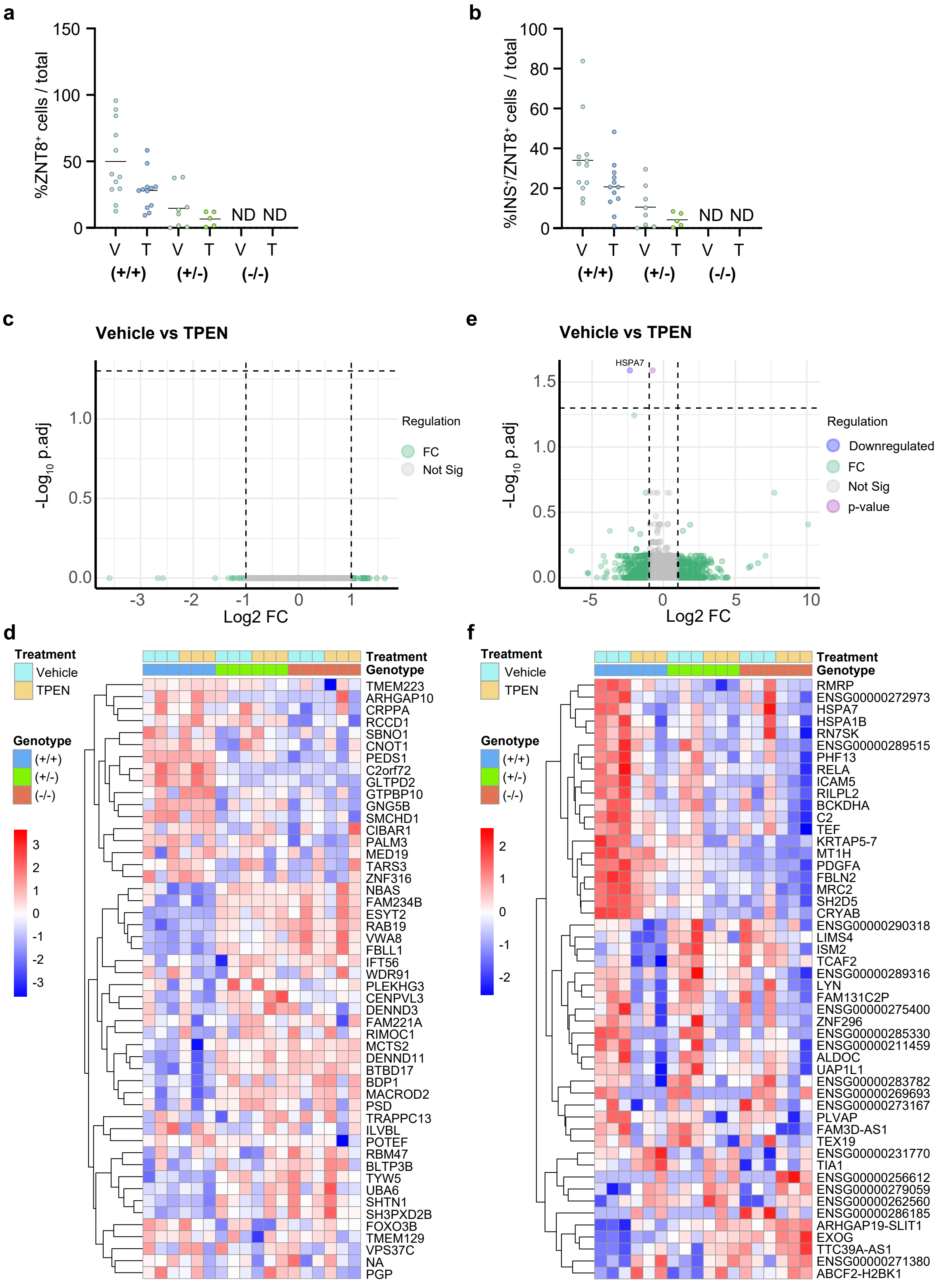
a
%ZNT8⁺ cells / total
150
100
50
0
ND ND
V T V T V T
(+/+) (+/-) (-/-)
b
%INS⁺/ZNT8⁺ cells / total
100
80
60
40
20
0
ND ND
V T V T V T
(+/+) (+/-) (-/-)
c
Vehicle vs TPEN
-Log10 p.adj
Log2 FC
Regulation
FC
Not Sig
e
Vehicle vs TPEN
HSPA7
-Log10 p.adj
Log2 FC
Regulation
Downregulated
FC
Not Sig
p-value
d
Treatment
Vehicle
TPEN
Genotype
(+/+)
(+/-)
(-/-)
TMEM223
ARHGAP10
CRPPA
RCCD1
SBNO1
CNOT1
PEDS1
C2orf72
GLTPD2
GTPBP10
GNG5B
SMCHD1
CIBAR1
PALM3
MED19
TARS3
ZNF316
NBAS
FAM234B
ESYT2
RAB19
VWA8
FBLL1
IFT56
WDR91
PLEKHG3
CENPVL3
DENND3
FAM221A
RIMOC1
MCTS2
DENND11
BTBD17
BDP1
MACROD2
PSD
TRAPPC13
ILVBL
POTEF
RBM47
BLTP3B
TYW5
UBA6
SHTN1
SH3PXD2B
FOXO3B
TMEM129
VPS37C
NA
PGP
f
Treatment
Vehicle
TPEN
Genotype
(+/+)
(+/-)
(-/-)
RMRP
ENSG00000272973
HSPA7
HSPA1B
RN7SK
ENSG00000289515
PHF13
RELA
ICAM5
RILPL2
BCKDHA
C2
TEF
KRTAP5-7
MT1H
PDGFA
FBLN2
MRC2
SH2D5
CRYAB
ENSG00000290318
LIMS4
ISM2
TCAF2
ENSG00000289316
LYN
FAM131C2P
ENSG00000275400
ZNF296
ENSG00000285330
ENSG00000211459
ALDOC
UAP1L1
ENSG00000283782
ENSG00000269693
ENSG00000273167
PLVAP
FAM3D-AS1
TEX19
ENSG00000231770
TIA1
ENSG00000256612
ENSG00000279059
ENSG00000262560
ENSG00000286185
ARHGAP19-SLIT1
EXOG
TTC39A-AS1
ENSG00000271380
ABCF2-H2BK1

**Supplementary Figure 1.** Zinc depletion has mild transcriptomic and proteomic effects. (**a-b**) Immunofluorescent image quantification of ZNT8$^{+}$ cells and INS$^{+}$/ZNT8$^{+}$ double positive cells of R138X$^{+/+}$, R138X$^{+/-}$ and R138X$^{-/-}$ islet-like clusters. (**c**) Volcano plots showing differential protein expressions between vehicle and TPEN treatment, while accounting the genotype (TPEN 1 μM, n = 3 independent differentiations). (**d**) Hierarchical clustering representing the top 50 most altered proteins between vehicle and TPEN treatment, as revealed by limma analysis. (**e**) Volcano plots showing differential gene expressions between vehicle and TPEN treatment while accounting the genotype (TPEN 1 μM, n = 3 independent differentiations). (**f**) Hierarchical clustering representing the top 50 most altered genes between vehicle and TPEN treatment, as revealed by DESeq analysis. Data were clustered by Ward's clustering with the Euclidean distance in R. Heatmap columns represent each genotype, in vehicle or TPEN conditions, and rows represent dysregulated proteins.

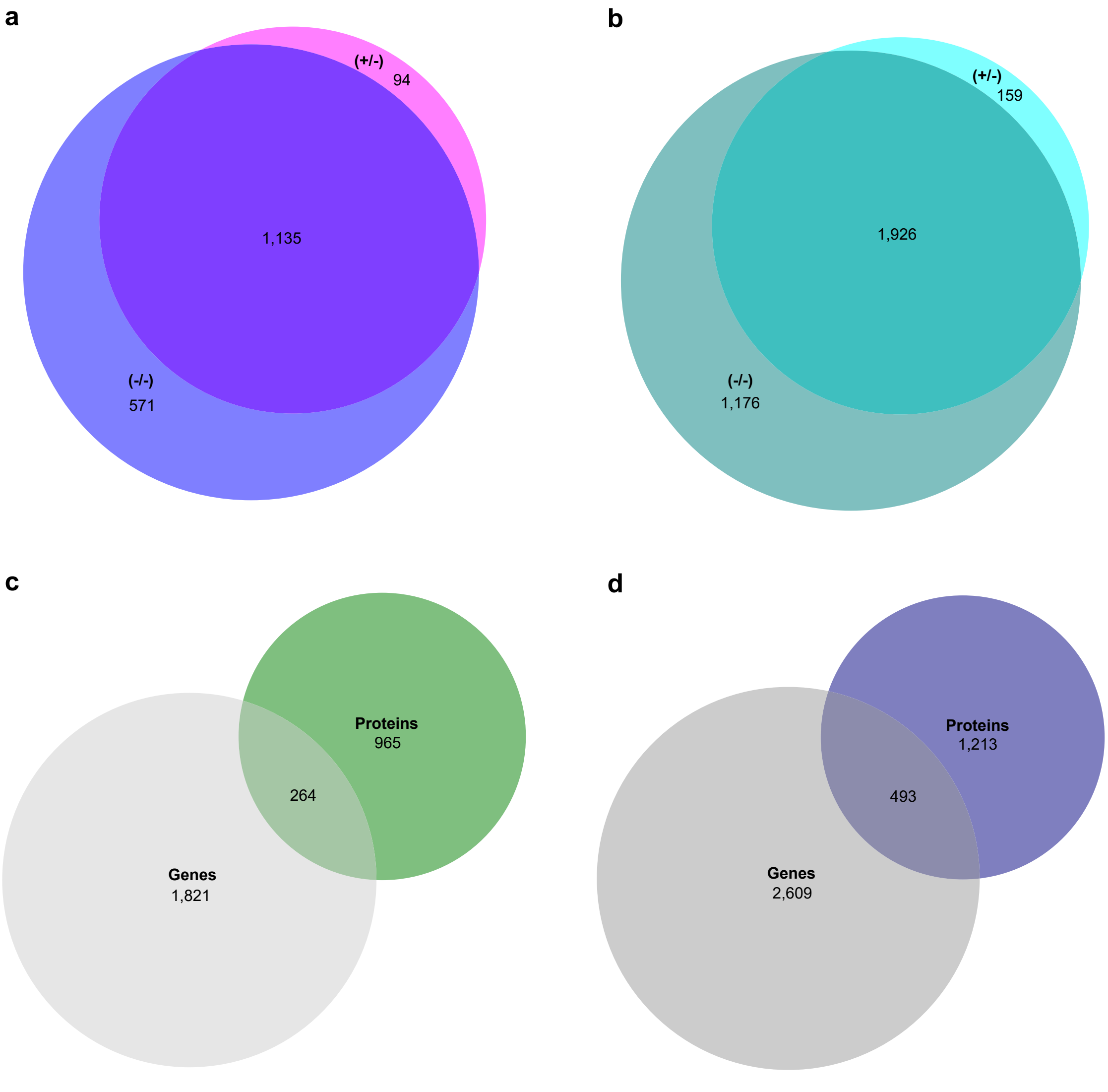
a
(+/-)
94
1,135
(-/-)
571
b
(+/-)
159
1,926
(-/-)
1,176
c
Proteins
965
264
Genes
1,821
d
Proteins
1,213
493
Genes
2,609

**Supplementary Figure 2.** (**a**) Venn diagram illustrating the overlap of significantly regulated proteins between R138X$^{+/-}$ and R138X$^{-/-}$ clusters based on proteomic analysis. Differential expression was defined as $log_2FC>1$ or <-1 and adj. p-value<0.05 by limma analysis. (**b**) Venn diagram illustrating the overlap of significantly regulated genes between R138X$^{+/-}$ and R138X$^{-/-}$ clusters based on transcriptomic analysis. Differential expression was defined as $log_2FC>1$ or <-1 and adj. p-value<0.05 by DESeq analysis. (**c**) Venn diagram illustrating the overlap of significantly regulated genes and proteins in R138X$^{+/-}$ proteomic and transcriptomic analysis. Differential expression was defined as $log_2FC>1$ or <-1 and adj. p-value<0.05. (**d**) Venn diagram illustrating the overlap of significantly regulated genes and proteins in R138X$^{-/-}$ proteomic and transcriptomic analysis. Differential expression was defined as $log_2FC>1$ or <-1 and adj. p-value<0.05. Numbers represent the number of proteins and genes in each category.

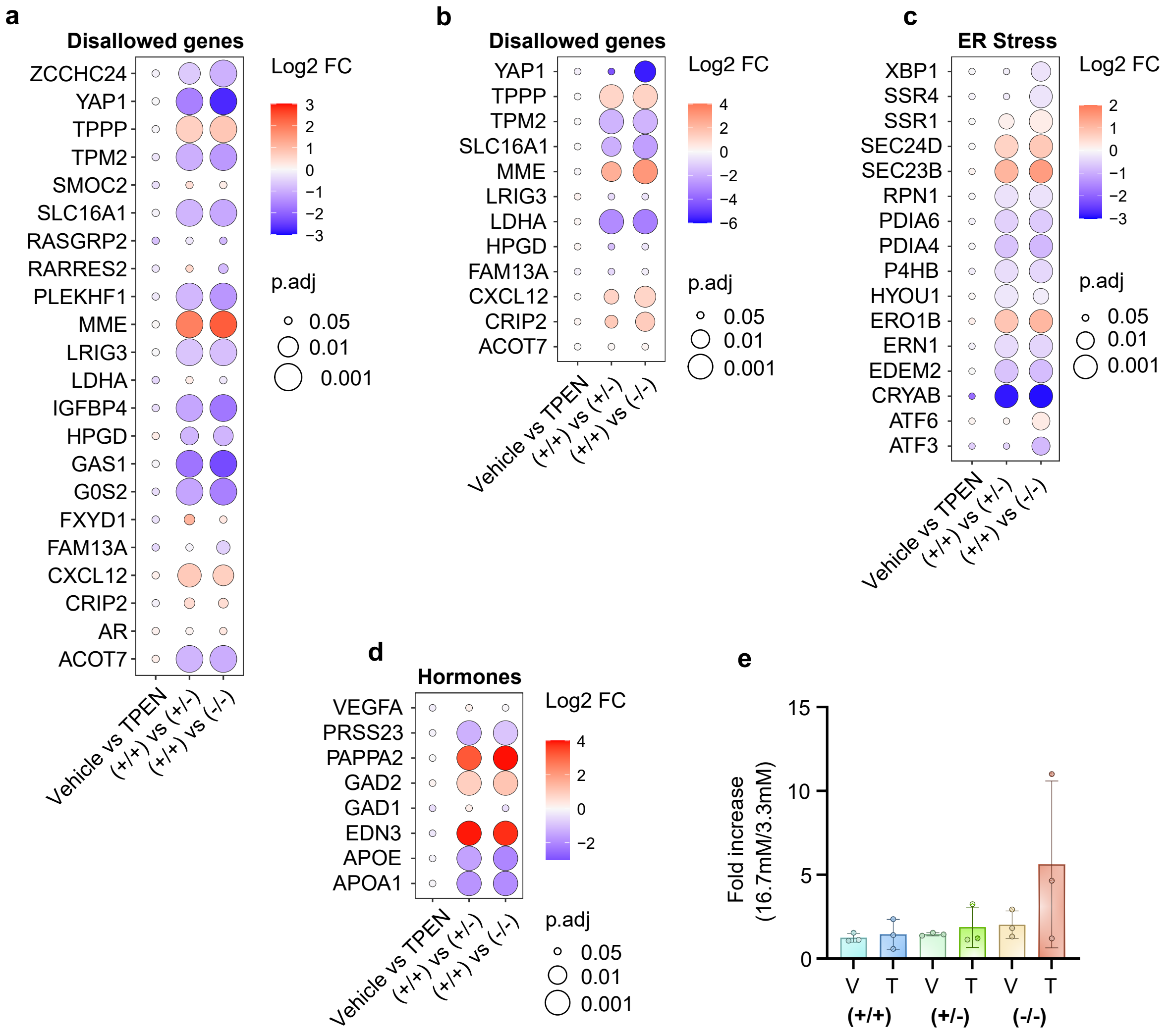

a
Disallowed genes
ZCCHC24
YAP1
TPPP
TPM2
SMOC2
SLC16A1
RASGRP2
RARRES2
PLEKHF1
MME
LRIG3
LDHA
IGFBP4
HPGD
GAS1
G0S2
FXYD1
FAM13A
CXCL12
CRIP2
AR
ACOT7
Vehicle vs TPEN
(+/+) vs (+/-)
(+/+) vs (-/-)
Log2 FC
p.adj
0.05
0.01
0.001
b
Disallowed genes
YAP1
TPPP
TPM2
SLC16A1
MME
LRIG3
LDHA
HPGD
FAM13A
CXCL12
CRIP2
ACOT7
c
ER Stress
XBP1
SSR4
SSR1
SEC24D
SEC23B
RPN1
PDIA6
PDIA4
P4HB
HYOU1
ERO1B
ERN1
EDEM2
CRYAB
ATF6
ATF3
d
Hormones
VEGFA
PRSS23
PAPPA2
GAD2
GAD1
EDN3
APOE
APOA1
e
Fold increase (16.7mM/3.3mM)
V
T
(+/+)
(+/-)
(-/-)

**Supplementary Figure 3.** The absence of ZnT8 disrupted genes and proteins associated with islet functions and cellular stress apoptosis. (**a**) Expression levels of disallowed genes. The dot colors represent the $log_2$FC and the dot size represents the adjusted p-value, as revealed by the DESeq analysis of the transcriptomic data. The treatment effect was evaluated while accounting the genotype (Vehicle vs TPEN). The genotype effect was evaluated while accounting zinc depletion, and comparison were made between R138X$^{+/+}$ and R138X$^{+/-}$ clusters ((+/+) vs (+/-)) and between R138X$^{+/+}$ and R138X$^{-/-}$ clusters ((+/+) vs (-/-)). (**b**) Protein expression levels of disallowed genes, as revealed by limma analysis of the proteomic data. Similar to transcriptomic results, the treatment effect was evaluated while accounting the genotype (Vehicle vs TPEN). The genotype effect was evaluated while accounting zinc depletion, and comparison were made between R138X$^{+/+}$ and R138X$^{+/-}$ clusters ((+/+) vs (+/-)) and between R138X$^{+/+}$ and R138X$^{-/-}$ clusters ((+/+) vs (-/-)). (**c**) Expression levels of genes related to endoplasmic reticulum stress. (**d**) Expression levels of hormone related genes. (**e**) Fold increase of insulin release at high glucose (16.7 mM) over basal glucose (3.3 mM) from R138X$^{+/+}$, R138X$^{+/-}$ and R138X$^{-/-}$ islet-like clusters after 27 days of differentiation and 48h of TPEN treatment (1 µM, n = 3 independent differentiations). One-way ANOVA followed by Tukey post hoc test were used to analyze differences groups and data are represented as mean ± SD.

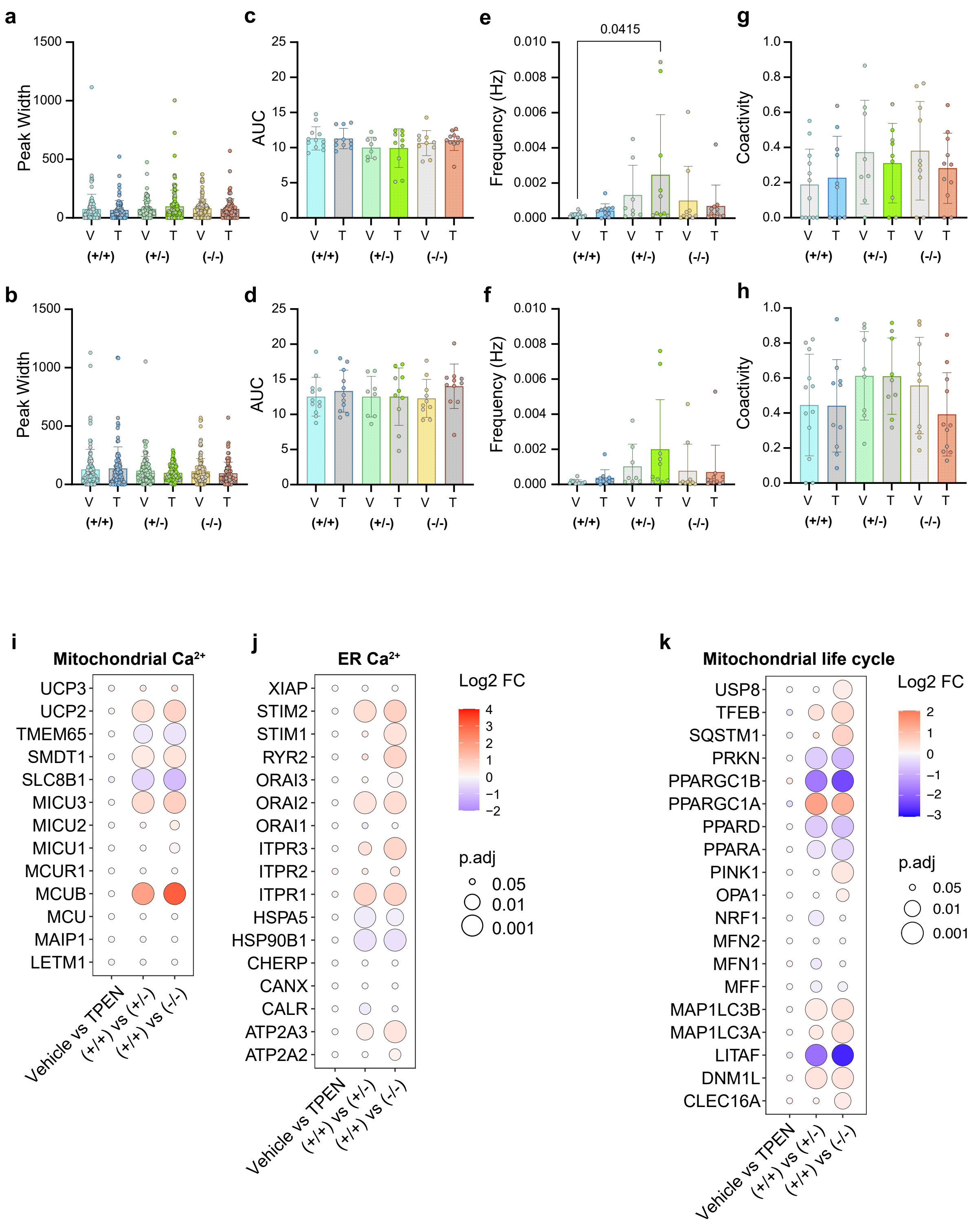

a
Peak Width
1500
1000
500
0
V
T
(+/+)
(+/-)
(-/-)
c
AUC
25
20
15
10
5
e
0.0415
Frequency (Hz)
0.010
0.008
0.006
0.004
0.002
0.000
g
Coactivity
1.0
0.8
0.6
0.4
0.2
0.0
b
d
f
h
i
Mitochondrial Ca²⁺
UCP3
UCP2
TMEM65
SMDT1
SLC8B1
MICU3
MICU2
MICU1
MCUR1
MCUB
MCU
MAIP1
LETM1
Vehicle vs TPEN
(+/+) vs (+/-)
(+/+) vs (-/-)
j
ER Ca²⁺
XIAP
STIM2
STIM1
RYR2
ORAI3
ORAI2
ORAI1
ITPR3
ITPR2
ITPR1
HSPA5
HSP90B1
CHERP
CANX
CALR
ATP2A3
ATP2A2
Log2 FC
4
3
2
1
0
−1
−2
p.adj
0.05
0.01
0.001
k
Mitochondrial life cycle
USP8
TFEB
SQSTM1
PRKN
PPARGC1B
PPARGC1A
PPARD
PPARA
PINK1
OPA1
NRF1
MFN2
MFN1
MFF
MAP1LC3B
MAP1LC3A
LITAF
DNM1L
CLEC16A
−3

**Supplementary Figure 4.** Zinc depletion and ZnT8 absence alter intracellular calcium signaling in islet-like clusters. (**a-b**) Corresponding $Ca^{2+}$ peak width at 11G (**a**) and 16.7G (**b**) from all individual cells across all analyzed islets. (**c-d**) Corresponding $Ca^{2+}$ area under the curve (AUC) at 11G (**c**) and 16.7G (**d**) from all individual islets. (**e-f**) Corresponding $Ca^{2+}$ oscillation frequencies at 11G (**e**) and 16.7G (**f**) from all individual islets. (**g-h**) Coactivity representing the fraction of time points where two (or more) cells were active at the same time at 11G (**g**) and 16.7G (**h**). Data are presented as mean ± SD. One-way ANOVA followed by Tukey post hoc test were used to analyze differences groups and data are represented as mean ± SD. (**i-j**) Expression levels of genes related to mitochondrial (**i**) and endoplasmic reticulum (**j**) $Ca^{2+}$ signaling. The dot colors represent the $log_2FC$ and the dot size represents the adjusted p-value, as revealed by the DESeq analysis of the transcriptomic data. The treatment effect was evaluated while accounting the genotype (Vehicle vs TPEN). The genotype effect was evaluated while accounting zinc depletion, and comparison were made between $R138X^{+/+}$ and $R138X^{+/-}$ clusters ((+/+) vs (+/-)) and between $R138X^{+/+}$ and $R138X^{-/-}$ clusters ((+/+) vs (-/-)). (**k**) Expression levels of genes related to mitochondrial life cycle.

**Movie S1.** $Ca^{2+}$ imaging in R138X$^{+/+}$ islet in vehicle condition.

**Movie S2.** $Ca^{2+}$ imaging in R138X$^{+/+}$ islet in TPEN condition.

**Movie S3.** $Ca^{2+}$ imaging in R138X$^{+/-}$ islet in vehicle condition.

**Movie S4.** $Ca^{2+}$ imaging in R138X$^{+/-}$ islet in TPEN condition.

**Movie S5.** $Ca^{2+}$ imaging in R138X$^{-/-}$ islet in vehicle condition.

**Movie S6.** $Ca^{2+}$ imaging in R138X$^{-/-}$ islet in TPEN condition.
